# A systematic review of the applications of Mendelian randomization assessing the causal relevance of the gut microbiome in human health and disease

**DOI:** 10.1101/2025.06.03.25328787

**Authors:** Charlie Hatcher, Alec McKinlay, Anna Bailward, Amy C. Dawes, David A Hughes, Dimitri J Pournaras, Laura J. Corbin, Lucy J. Goudswaard, Francesca Spiga, Kaitlin H. Wade

**Author notes:** **Corresponding author:** Dr Kaitlin Wade, Oakfield House, Oakfield Grove, BS8 2BN. contributed equally (Charlie Hatcher, Kaitlin H. Wade).

## Abstract

**Objectives:** To investigate the current application of Mendelian randomization (MR) in assessing the causal relevance of the gut microbiome in human health and evaluate the quality of these studies.

**Design:** Systematic review

**Data sources:** Ovid MEDLINE, Embase, Web of Science, bioRxiv and medRxiv were searched from inception to the 12^th^ of January 2023.

**Eligibility criteria:** Full-texts and conference abstracts of studies that conducted MR analyses to investigate the causal role of the gut microbiome on any outcome.

**Methods and analysis:** Two independent reviewers screened titles and abstracts, assessed full texts for eligibility, extracted data and assessed study quality. Extracted data included information on authors, hypothesis/rationale, methodology used (including genetic instrumentation decisions and analyses), results and limitations. As no quality assessment tool currently exists for MR studies, the quality of each study was assessed using a series of questions adapted from two previous systematic reviews of MR studies and a comparison with the STROBE-MR guidelines. Results were narratively synthesized, and meta-analyses were conducted, where possible, if the exposure and outcome were comparable (including definition and units) and data sources were appropriately independent across studies.

**Results:** Of the 463 records identified, 66 were eligible for inclusion. We identified 48,082 individual MR estimates of the relationship between 612 gut microbial traits (defined by relative abundance, presence vs. absence or functional pathway) and 905 health outcomes including those categorized into autoimmunity, behaviour, cancer, prescription drug usage, immunity, inflammation, longevity, medical procedures, metabolic health, nutrition, pain, sexual and reproductive health, and diseases of several organs and systems. According to the quality assessment, all studies were judged to be of poor quality, due to the inappropriate application of MR – specifically, instrument selection, exposure and outcome definition, choice of analytical methodology, assessment of reverse causation and replication – and lack of transparent reporting of findings. Therefore, meta-analysis across studies was largely impossible.

**Conclusions:** Whilst there has been growth in the application of MR to understand the causal role of the microbiome in human health, these studies fail to appropriately apply the method and transparently report findings. Further, our systematic review provides evidence of an unmet requirement for careful examination and interpretation of derived causal estimates. Here, we make recommendations for the improvement of applications of MR to the microbiome going forward.

**Study registration:** https://www.crd.york.ac.uk/prospero/display_record.php?RecordID=314055

**SUMMARY BOX:** *What is already known on this topic:*

- Mendelian randomization (MR) is increasingly used to assess the causal relevance of the gut microbiome in human health.
- Concerns exist regarding the methodological quality, validity of MR studies in this context and, thus, the level of misinformation entering the public domain.
- There is a requirement to evaluate the application and reporting quality of these studies.

*What this study adds:*

- Our findings show that most MR studies investigating the gut microbiome and health outcomes are of poor quality due to methodological flaws and inadequate reporting.
- Our study highlights the urgent need for improved study design, rigorous

## INTRODUCTION

The human gut microbiome is a complex system of microorganisms that aids digestion, provides protection against pathogens, improves immune response and creates essential metabolites. Over the past few decades, interest in the gut microbiome as a possible determinant or predictor of health outcomes has grown substantially^2^. Whilst evidence from model organisms and human studies support such relationships, the causal role of the gut microbiome in health and disease remains ambiguous, with many inconsistencies frequently reported in the literature^3^. This is particularly alarming given the growing market for commercial initiatives targeting the microbiome as a consumer-driven intervention for improving health. Few findings in model organisms have been translated to humans and there remains an active debate as to whether animal models of the human gut microbiome are translatable to humans for all scientific questions and scenarios^4,5^. Human studies in this context are largely observational and cross-sectional in nature, often suffering from confounding (e.g., dietary factors and antibiotics), bias and reverse causation. As such they are unable to confidently provide evidence for or against causality. In addition, most studies of the gut microbiome and specific diseases tend to focus on outcomes after diagnosis (e.g., treatment effects or prognosis) rather than the onset of disease; thus, limiting their ability to provide evidence relating to disease prevention.

In lieu of conducting long-term, expensive and potentially unfeasible randomized controlled trials (RCTs) of an exposure - such as microbiota perturbations - on disease outcomes, Mendelian randomization (MR) is an established alternative approach that aims to improve causal inference in epidemiological relationships^1,6,7^. Relying on the random and fixed nature of germline genetic variation, MR uses genetic variants (usually single nucleotide polymorphisms [SNPs]) as “proxy” measures or “instrumental variables” (IVs) for an exposure of interest (e.g., the gut microbiome). This approach assesses the existence, direction and magnitude of a causal effect between that exposure and an outcome, provided that key assumptions are met (Figure 1). The method can be applied within individual-level data (commonly denoted as “one-sample MR” or “MR with individual-level data”) or, more recently with the advent of genome-wide association studies (GWASs) and accompanying published results describing the relationships between SNPs and traits, using summary-level data (commonly denoted as “two-sample MR” or “MR with summary-level data”; Box 1)^6,8^.

**Figure 1.**
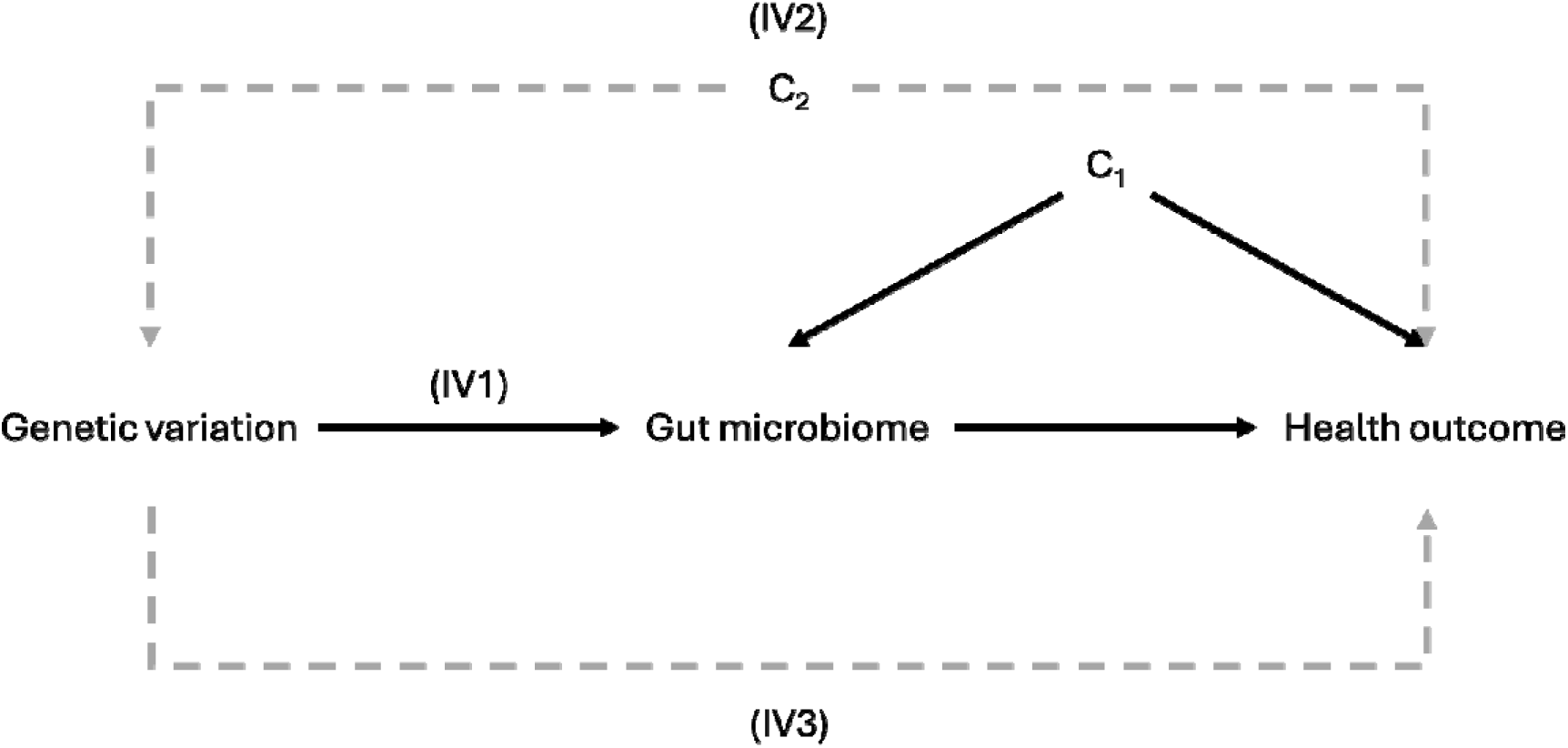
The core assumptions of Mendelian randomization (MR) IV = instrumental variable; MR = Mendelian randomization. MR relies on three core assumptions to test for the presence of a causal effect: (IV1) the genetic variant(s), typically single nucleotide polymorphisms (SNPs) must be robustly and strongly related to the exposure of interest (here, the gut microbiome), usually ensured by using SNPs that reach a p-value threshold accounting for the number of independent tests undertaken when identifying such SNPs (e.g., p<5e-08 in a conventional genome-wide association study); (IV2) there must be no common causes (i.e., confounders) of the SNP(s) being used as an instrument for the exposure and the outcome (C_2_) – such confounders will be those at the population-level (e.g., any population structure such as ancestry, dynastic or intergenerational effects and assortative mating) and are often unlikely to be the same confounders of the exposure and outcome association (C_1_); and (IV3) there must be no effect of the SNP(s) on the outcome independent of the exposure – an assumption that is also likely violated if instruments are selected using a lenient p-value threshold. These assumptions are commonly referred to as the “relevance” (IV1), “independence” (IV2) and “exclusion restriction” or “no horizontal pleiotropy” (IV3) assumptions. A further set of additional assumptions (known as the “homogeneity”, “monotonicity” and “no effect modification” assumptions) are also required to estimate the direction and magnitude of a causal effect. Full details of MR and its assumptions can be found on the MR Dictionary (https://mr-dictionary.mrcieu.ac.uk/).

Given its properties and relative ease of application, MR has been increasingly applied to understand the relevance of the human gut microbiome in several health outcomes^3,9,10^. This is particularly true when using summary-level data, a practice that has become more common with the growth of gut microbiome GWASs (mGWASs) and the development of tools like MR-Base^11^. However, an increasingly frequent, detrimental and reputationally damaging outcome of easily accessible data and analysis platforms is the preponderance of poorly conducted MR studies that sometimes present biologically implausible findings^12,13^. Particularly relevant to applying MR to gut microbial phenotypes, there are several complex challenges when using host (i.e., human) genetic variation as IVs for the microbiome (i.e., a different organism to the host) that are rarely appropriately considered. The main challenge is the substantial lack of robustly associated host genetic variants replicated across mGWASs (i.e., at a genome-wide p-value threshold of 1×10^−8^) and understanding of the mechanisms by which these variants influence the microbiome. Within this context, the recurrent unjustifiable relaxations of IV selection criteria (i.e., p-value thresholds) likely lead to violations of core MR assumptions (Figure 1). Therefore, there are concerns about the quality of current applications of MR in assessing the causal relevance of the gut microbiome in human health and disease. Here, we systematically reviewed the application of MR to the gut microbiome, summarising the evidence provided and assessing study quality.

## METHODS

The protocol for this systematic review was published on PROSPERO prior to starting the review (https://www.crd.york.ac.uk/prospero/display_record.php?RecordID=314055).

### Eligibility criteria

A study was considered eligible if MR methodology was used to investigate the causal role of the gut microbiome on any outcome. Participants were those whose data were used in these MR analyses. The exposure of interest was any specified measure of intestinal microbiota, specified in the search strategy by a variety of terms and synonyms. As we were interested in the causal effect estimates between the gut microbiome and any outcome in an MR framework, there were no controls or comparators, and no outcome specified. The record must also have been written in English and be available in full text. Where studies used SNPs as IVs or measured the direct association between genetic variation (relating to the gut microbiome) and an outcome, these were included even though the method was not explicitly referred to as MR (provided they met the other inclusion criteria). Additionally, if a record focused on the gut microbiome alongside other exposures, the record was included and only the information for the microbial exposures were reported, if available, independent from the other exposures. Reviews and commentaries obtained through the searches were also included in the title/abstract and full-text screening phases, where their reference lists were screened as an additional source of relevant records and included if they met the other inclusion criteria.

### Data sources and search strategy

Search strategies were developed by one reviewer (FS) with the assistance of an information specialist using key words including the gut microbiome, MR methodology and their respective synonyms (see Supplementary Materials). EMBASE, Ovid MEDLINE and Web of Science bibliographic databases alongside the pre-print servers, bioRxiv and medRxiv, were searched using a combination of free-text and controlled vocabulary terms from inception until the 12^th^ of January 2023.

### Record selection

Records obtained through searching the databases described above were imported into EndNote (version 20.2; Clarivate) and de-duplication was performed using EndNote’s in-built software^14^. Titles and abstracts of all remaining, de-duplicated records were uploaded to Rayyan^15^, screened independently by two reviewers (CH and KHW) to check possible alignment with the pre-defined inclusion criteria (see below) and conflicts were resolved through discussion between the reviewers. Where a pre-print (i.e., published on bioRxiv or medRxiv) had been published and captured in the search of searched databases both records were compared to check whether the same information was presented. If this was the case, the published version was included over the pre-print version (which was excluded), otherwise, both records were included. The full texts of these included records were then evaluated for eligibility by the same two reviewers independently, keeping only those records that met the above criteria.

### Data extraction

Study meta-data and summary statistics of the effect of each gut microbial trait on any health outcome from the main MR analysis (which could be either one- or two-sample methodology) were extracted from each study using a pre-piloted data extraction form. This form was adapted from our previous systematic review on the effects of adiposity on health outcomes^16^ and based on the STROBE-MR guidelines^17,18^. Meta-data were extracted by one reviewer (CH or KHW) and checked by the other reviewer. Summary statistics of the effect estimates were extracted independently by two reviewers (CH and KHW). Any discrepancies were resolved through discussion. The following data were extracted from each study and compiled into a single spreadsheet (see Supplementary File 1):

- **Paper identifiers** (title, publication details, first and corresponding authors’ information, hypotheses and rationale)
- **Exposure and outcome information** (exposure and outcome measurements, definitions and units)
- **Data sources** (GWASs or studies from which genetic, exposure and outcome information were sourced and their demographics, sample size and ancestry as well as genotyping and imputation approach, covariate adjustment, determination of linkage disequilibrium (LD), test for Hardy-Weinburg equilibrium and number of SNPs identified)
- **Instrumentation information** (p-value threshold(s) used to select IVs and comparison with GWAS results, use of proxy SNPs, harmonization processes, number of IVs used and their strength)
- **Study design and methodology** (whether the analyses used were one- or two-sample MR, the main analysis method, whether any corrections for multiple testing were applied and software used)
- **Sensitivity analyses** (methods used to test for violations of MR assumptions, predominantly the exclusion restriction or “no horizontal pleiotropy” assumption)
- **Power and replication** (whether the studies reported power calculations and whether replication was sought)
- **Summary statistics of MR estimates** (effect estimates, standard errors and/or confidence intervals (CIs) and p-values, extracted in duplicate)
- **Discussion of study** (specifically, the limitations described including likelihood of violations of MR assumptions through horizontal pleiotropy and population stratification, and statistical power).

When a study used data from a public source (e.g., the GWAS from which the exposure and outcome data were obtained in a two-sample MR), we checked that the information provided in the study was consistent with the original source material. Each exposure was categorized based on phenotypic characteristics (e.g., bacterial measurement or pathway). Similarly, for each outcome, we used a two-level categorization, grouping similar outcomes together based on their clinical characteristics, whilst still maintaining information about the type of outcome being analysed. This categorization process of both the exposure and outcome allowed more efficient analyses. For example, “Alzheimer’s disease” was categorized in the “Neurogenerative diseases” group (first level) and “Brain” group (second level, representing a higher-order categorisation of all outcomes). Where relevant data were not reported or it was not possible to decipher the information provided by the authors, the data extraction spreadsheet was left blank (or an equivalent to “NA” was entered instead).

For efficient data synthesis, extracted information was checked and harmonized such that the format of all entries across studies was consistent. Trait names were harmonized, favouring the most informative entry. For example, suppose one study reported the effect of an exposure (“G_Bifidobacterium_RNT”) on an outcome (“BMI”) and another study reported an effect of the same exposure (that was instead called “Bifidobacterium (Genus)” on the same outcome (that was instead called “Body mass index”). After harmonization, both studies would be recorded with the exposure specified as “Bifidobacterium (Genus)” and the outcome as “Body mass index”. A column was then added to provide any additional relevant details (e.g., “Abundance (SD)” to clarify the measure and unit of the exposure were rank normal transformed relative abundance values).

If authors used a pre-existing GWAS for a particular exposure or outcome, but did not provide either the sample size or enough information to know precisely which GWAS was used, the data source was listed as an “Unknown GWAS”. Equally, if the authors provided a sample size of the GWAS used but that sample size did not match the referenced GWAS, this was also listed as an “Unknown GWAS”. Additionally, if a study had specified a GWAS identifier as the source of outcome data (as defined by MR-Base^11^), rather than the original GWAS itself, the original GWAS was found and this information was added to the data extraction spreadsheet.

### Meta-analysis methodology

MR estimates were meta-analysed if they met a series of criteria (defined previously^16^). MR estimates assessing the same exposure-outcome association were meta-analysed if the exposure (and its units) were known and the same, the outcome (and its units) were known and the same, and the sources of outcome data were independent across MR estimates (Supplementary Figure 1). The risk of bias of using the same exposure data across MR estimates being meta-analysed is low; therefore, this was not one of the criteria for meta-analysis^19^. Meta-analyses were conducted using estimates presented within-study (e.g., where authors had replicated their analyses within the same paper) and between-studies (e.g., where results were comparable across studies) and met the meta-analysis criteria described above.

In line with our previous work^16^, an inverse-variance weighted random-effects model was used to meta-analyse effect estimates with accompanying standard errors using the *meta* package (version 8.0-2) in R and the function *metagen*^20^. If standard errors were not available, these were calculated using available effect estimates and/or confidence intervals (CIs) and p-values provided. For continuous outcomes, effect estimates represent the mean difference of each outcome on the original scale (e.g., mean standard deviation (SD) difference) with each exposure. For binary outcomes, effect estimates represent the difference in risk based on the relevant summary method (e.g., odds or risk ratio). Characteristics of all included studies were summarised, where only meta-analysed MR estimates were reported and discussed (i.e., we did not narratively synthesize the results of studies that were not eligible for meta-analysis).

### Quality assessment

As there is currently no risk of bias or quality assessment tool to assess the quality of MR studies, the quality of each study was assessed using a series of questions adapted from two previous systematic reviews of MR studies (including our previous publication assessing the role of adiposity on health outcomes)^16,21^ and a comparison with the STROBE-MR guidelines^17,18^ (see Supplementary File 2). Study quality was assessed on a 3-point scale (i.e., low = 3, moderate = 2, high = 1 quality) across 19 questions (also referred as “items”) each addressing a specific potential source of bias: IV selection, collider bias, selection of participants and overlap between samples used (in a two-sample MR analysis), statistical overfitting, harmonization processes, and choice (and execution) of main and sensitivity analyses. The quality assessment (QA) tool also queried whether authors appropriately tested for violations of MR assumptions including instrument strength, genetic confounding and horizontal pleiotropy (Figure 1). Additionally, the QA tool queried whether authors had tested for possible reverse causation in an MR framework (i.e., whereby the “outcome” influences the “exposure”) and had replicated their findings. Lastly, the QA tool assessed whether authors defined the IVs, exposure and outcome adequately, and described the methods and results transparently and reproducibly. As each item of the QA tool was equally important, any study that had a “low” quality (i.e., a score of 3) judgment across any of the 19 questions was considered to be of low quality overall. The QA tool was not used as a prerequisite for inclusion/exclusion in the meta-analyses; rather, it was used to supplement the meta-analyses and aid interpretation.

As an additional assessment of quality across studies, MR estimates of the same exposure-outcome associations (i.e., where authors had used the same data sources and methodology and were objectively repeating the same analyses) were compared.

### Patient and Public Involvement

No patients or members of the public were involved in this research.

## RESULTS

### Search results and included studies

Of the 463 records identified by the searches, 112 met the pre-defined inclusion criteria for full-text evaluation (Figure 2). Of these, 47 records were excluded due to: (i) having no MR estimate of the gut microbiome as an exposure on an outcome (n=27), (ii) being a correspondence, editorial or review with no additional MR estimate (n=9) or (iii) being a duplicate pre-printed version of a published paper (n=11). Screening of the reference lists of correspondences, editorials and literature reviews did not identify any additional eligible study. After completing full-text screening, we also checked whether any pre-prints had since been published (i.e., that weren’t captured in the initial searches), which was the case for one record. Therefore, 66 studies were included (of which 56 were published in peer-review journals, seven were conference abstracts and three were unpublished pre-prints. Characteristics of included studies are reported in Supplementary Table 1.

**Figure 2.**
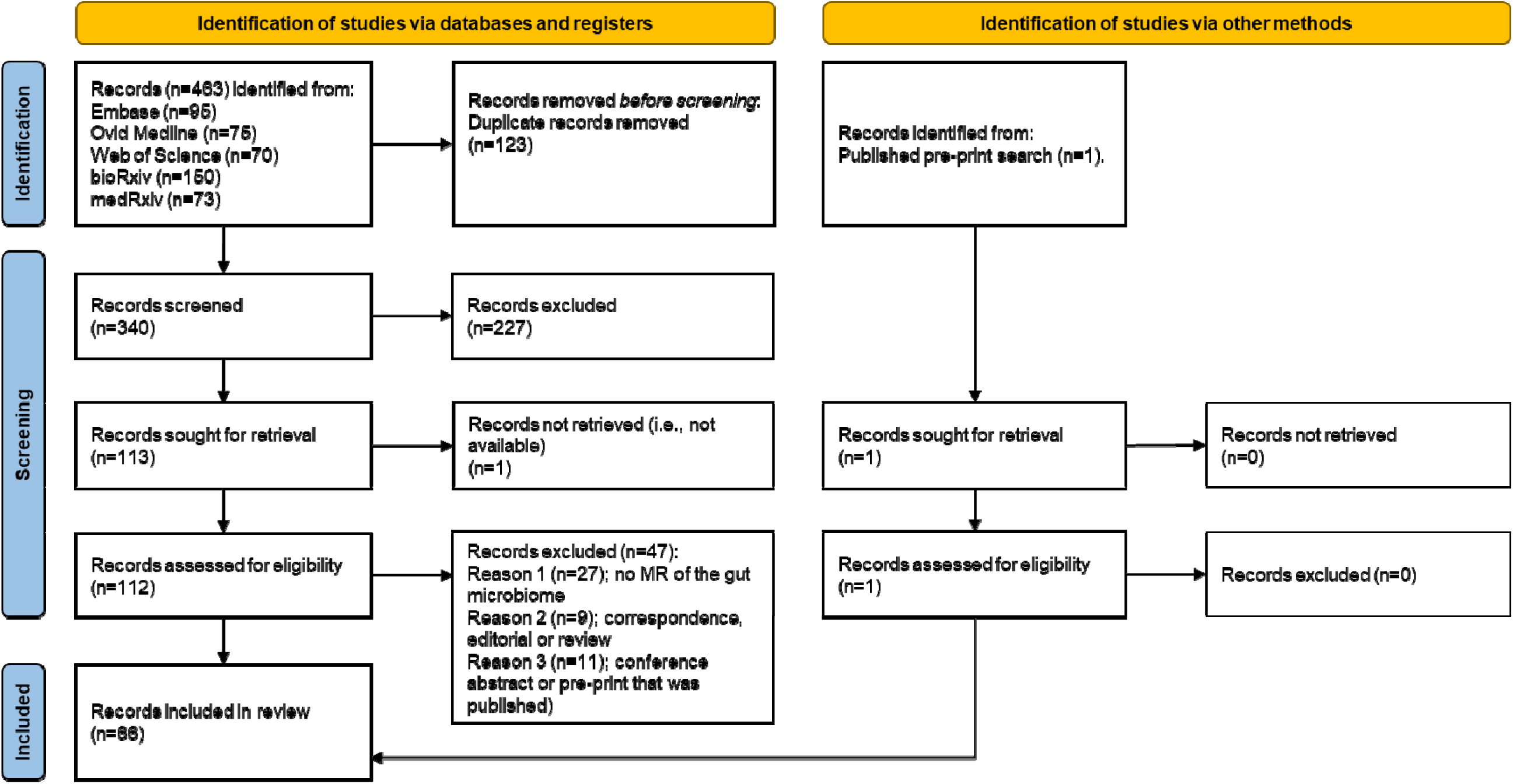
The Preferred Reporting Items for Systematic reviews and Meta-Analyses (PRISMA) flow chart of included studies. *From:* Page MJ, McKenzie JE, Bossuyt PM, Boutron I, Hoffmann TC, Mulrow CD, et al. The PRISMA 2020 statement: an updated guideline for reporting systematic reviews. BMJ 2021;372:n71. doi: 10.1136/bmj.n71. For more information, visit: http://www.prisma-statement.org/

### Characteristics of included studies

#### Journals and institutions

All included studies were published since 2018, with 52.5% published in 2022 and 28.8% published in 2021. Studies were published across 52 journals, with the most common being *Nature Genetics* (n=6; 9.1%), *medRxiv* (n=3; 4.6%) and *Frontiers in Microbiology* (n=3; 4.6%) and the most common publishing groups being *Frontiers* (n=15; 22.7%), followed by *Nature* (n=9; 13.6%) and *BMC* (n=4; 6.1%). Approximately 56% (n=37) of the 66 studies were published by first and/or corresponding authors from Chinese institutes, 10.6% (n=7) from institutes in the Netherlands, the same number (n=7; 10.6%) from the United Kingdom (UK) institutes and 9.1% (n=6) from institutes in the United States of America (USA).

#### Exposures and outcomes

Within the 66 studies, there were 48,082 individual MR estimates of the relationship between 612 unique gut microbial traits (defined by relative abundance, presence vs. absence or functional pathway; Figure 3) and 905 unique health outcomes including those categorized into autoimmunity, behaviours, cancer, prescription drug usage, immunity, inflammation, longevity, medical procedures, metabolic health, nutrition, pain, sexual and reproductive health, and diseases of several organs and systems (Figure 3). Most estimates reflected the effect of changes in the gut microbiome on outcomes relating to metabolic health (43.6%), nutrition (19.6%), the brain (11.7%) and autoimmunity (6.1%), though the occurrence of the outcomes categorised into the former two outcome groups were likely reflective of the high number of metabolites and nutritional variables (e.g., from food frequency questionnaires). A small proportion of outcomes (n=100; 0.2%) were not clear based on the information provided by the study authors and thus the outcome group was labelled as “Unknown”. The median number of MR estimates across the 66 studies was 22.5, with a mean of 728.5 and a minimum and maximum number of 1 and 17550, respectively.

**Figure 3.**
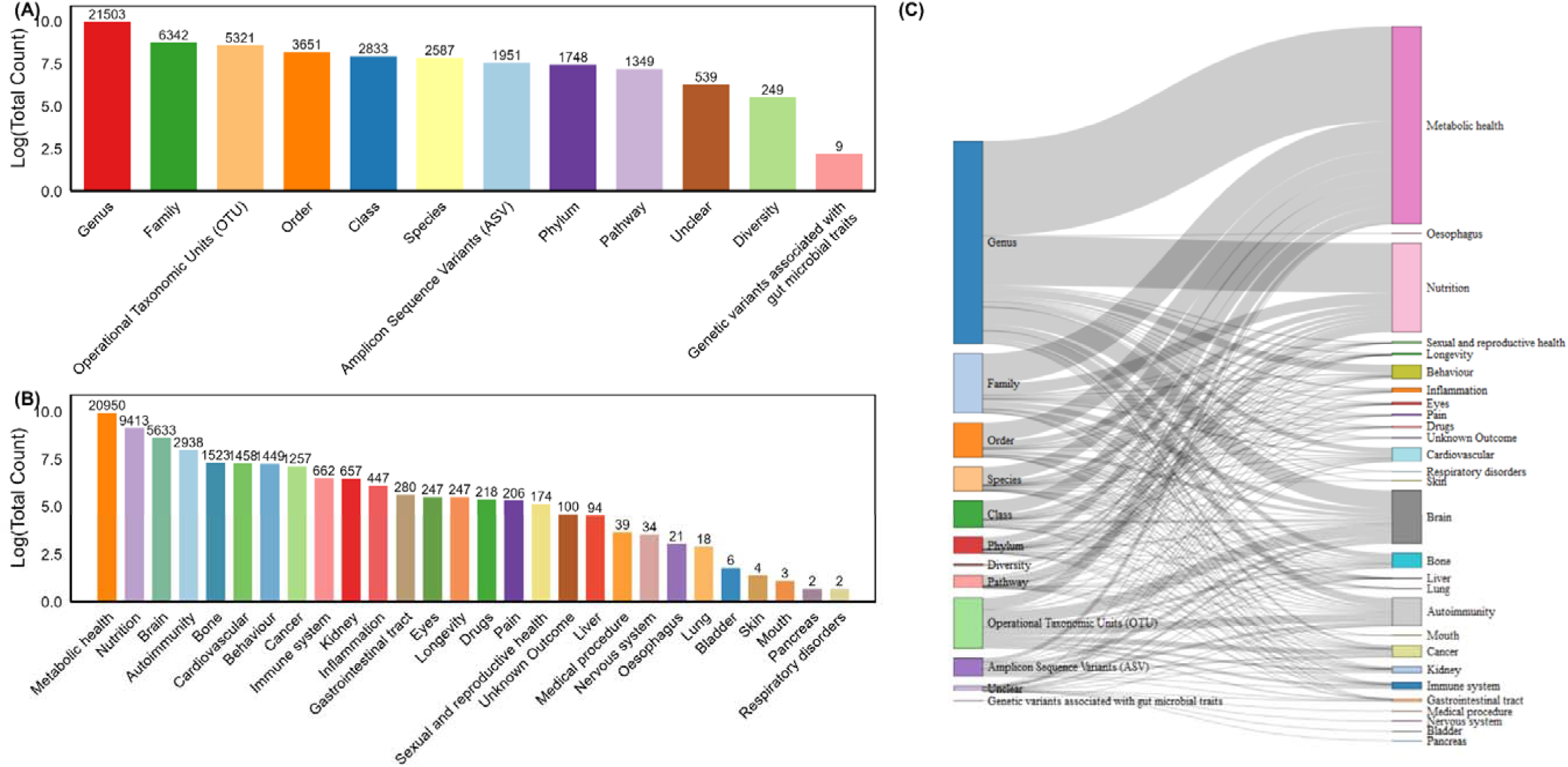
Visualisation of the types of exposures and outcomes observed across studies. (A) X-axis delineates each exposure category, analysed across all 66 studies, that were included in this review and the y-axis is the log_10_ total number of Mendelian randomization (MR) estimates for that exposure category on any outcome, where numbers atop each bar represent the absolute total number; (B) X-axis delineates each outcome group (i.e., categories of the types of outcomes) analysed across all 66 studies included in this review and the y-axis is the log_10_ total number of MR estimates for that outcome group given any number of possible microbial trait exposures, where numbers atop each bar represent the absolute total number; and (C) Sankey plot showing the relationships between each of the exposure and outcome groups across the 66 studies included in this review.

Over 99% of the gut microbial traits were measures of the abundance or presence / absence of certain bacteria (e.g., quantified via 16S ribosomal ribonucleic acid (rRNA) or metagenomic sequencing) and 0.1% of the gut microbial traits reflected changes in bacterial functional pathway (e.g., characterised by metagenomic sequencing). Specifically, most estimates described the effect of changes in bacterial genera (44.7% of all estimates), families (13.2%) or operational taxonomic units (OTUs; 11.1%) on various outcomes (Figure 3). A small proportion of estimates represented the effect of gut microbial exposures that were not clear based on the information provided by the study authors. This was either because they were described as “Unknown” (n=247) or “Unclassified” (n=1407), or because they were not defined (N=539; 1.1%) – i.e., carrying a broad label like a “Gut microbiome” phenotype or “functional units”. One study also reported the effect estimates of the associations between SNPs (associated with gut microbial traits) and the outcomes, rather than an MR estimate explicitly, representing nine of the 48,082 estimates.

#### MR framework and data sources for the gut microbiome

Almost all (99.9%) of the estimates across studies were generated using two-sample MR methodology and approximately 0.1% were generated using one-sample MR methodology. In one study^22^, the study design and methodology were not completely transparent as it reported using “one-sample MR” but two-sample MR analysis was described in the methods (Box 1). Of the 48,082 estimates, 52.4% (n=25,194) were generated using the MiBioGen mGWAS published by Kurilshikov *et al*.^10^ as the source of exposure (i.e., gut microbiome) information (i.e., the SNP-exposure association) for MR analyses, followed by the mGWASs published by Rühlemann *et al*.^23^ (n=9,602; 20%) and Hughes *et al*.^9^ (n=8,084; 16.8%). For a small proportion of estimates (n=66; 0.1%), the specific exposure data source was not reported.

Fourteen studies described mGWAS analyses that were performed to obtain associations between SNPs and the gut microbiome (with sample sizes ranging from 311 to 18,340 individuals) for MR analyses. Of these, six studies also used the same population sample as the source of SNP-outcome data (i.e., the MR analysis was performed in the same population sample within which the mGWAS was conducted), likely leading to overfitting and biasing effect estimates towards the null in the presence of weak instruments (Box 2).

Most studies (n=46; 69.7%) used one mGWAS source to identify SNPs associated with the exposure, as is expected given common practice in MR studies. In contrast, 12 studies identified and extracted SNPs from multiple studies for the purposes of bringing them together for one MR analysis. This is not best practice (Box 1, Box 2), unless authors explicitly make use of a discovery and replication study design or the mGWASs are fundamentally and directly comparable.

#### Meta-analysis

##### Within-study replication

Of the 66 studies, eight explicitly indicated that they had undertaken a replication^24–31^. However, when consulting the defined criteria for meta-analysis (i.e., that, across estimates, the exposure and unit are the same, the outcome and unit are the same and the outcome data sources are independent; Supplementary Figure 1), only four of these studies (Sanna *et al*.^24^, Liu *et al*.^26^, Liu *et al*.^25^ and Xu *et al*.^27^) included analyses that were truly replicated. The reasons that others were not considered as having undertaken a true replication were that they included (i) two exposure data sources (i.e., used to obtain the SNP-exposure associations) but not two independent outcome sources (i.e., for the SNP-outcome associations)^28^, (ii) two outcome data sources were claimed to be used but it was unclear what one of the data sources was and thus it was indeterminable whether the sources were truly independent^29^, (iii) two different outcome data sources but those sources comprised individuals of different populations (e.g., different ancestries)^30^ or (iv) explicitly non-independent outcome data sources^31^. There were no other studies that contained estimates that met the meta-analysis criteria.

Of the four studies that published replicated results, Sanna *et al*.^24^ undertook discovery and replication MR analyses assessing the role of the 245 microbiome features (including faecal short-chain fatty acid levels, unique taxa and pathways) and 17 obesity-related and glycaemic traits, including BMI and type 2 diabetes (T2D). Data from the GWAS analysis undertaken in the same study were used as the source of exposure-related summary statistics and data from outcome-specific GWAS were used as the discovery source for summary statistics for each outcome alongside UK Biobank as the replication data source. Focusing on the unique taxa and pathways analysed (as microbiome-related metabolites were not the focus of this review), only results for the pathway involved in 4-aminobutanoate (GABA) degradation (PWY-5022) were presented in the discovery analyses. Whilst there was evidence for a causal effect of PWY-5022 on several glucose- and insulin-related phenotypes, these were not present in UK Biobank. Analyses in UK Biobank also included five outcomes that were not analysed in the discovery data (body fat percentage, subcutaneous and visceral adipose tissue, waist-hip ratio (WHR; whereas the discovery analyses included WHR adjusted for BMI), and obesity). Therefore, the only phenotypes that were present in both analyses were BMI and T2D, using Locke *et al.* (for BMI)^32^ and Scott *et al*. (for T2D)^33^ in the discovery (however, note that there were no units explicitly provided for BMI in the replication dataset). In meta-analyses, there was little evidence for a causal effect of the PWY−5022 pathway on BMI and T2D, with the contributing effect estimates of the latter outcome being in opposite directions (see GitHub for relevant <u>figures</u> and <u>results</u>).

The Liu *et al*.^26^ study, undertook observational analyses and both one- and two-sample MR analyses of 500 microbial features with 112 metabolic traits using subsamples of a population cohort of Chinese individuals in which the GWAS analyses of both the exposure and outcome were undertaken, alongside two-sample MR analyses of these microbial features on 42 diseases and 59 quantitative traits in blood with Japan Biobank as the source of outcome data. Of all analyses undertaken, results from 18 associations between 13 gut microbial traits and nine different circulating metabolites were present in both the discovery and replication samples from the same population cohort. Of these, only four associations between four microbial traits and two metabolites were also present in Japan Biobank. Results provided some evidence that higher abundances of: (1) bacteria in the *Alistipes* genus lowered triglycerides and vitamin “VB5”, (2) bacteria in the *Faecalibacterium prausnitzii* species lowered selenium, (3) unclassified bacteria in the *Lachnospiraceae* family increased uric acid, (4) bacteria in the *Mobiluncus* genus and *Mobiluncus curtisii* species reduced alanine, (5) bacteria in the *Oscillibacter* genus reduced alanine and triglycerides, (6) bacteria in the *Pseudomonadales* order and *Salmonella* genus lowered 5−methyltetrahydrofolic acid and (7) bacteria in the *Streptococcus parasanguinis* species lowered strontium levels. Additionally, the study provided some evidence that (1) higher levels of the MF0004 pathway (involved in putrescine degradation) increased alanine and uric acid and lowered progesterone, (2) the MF0010 pathway (involved in sucrose degradation) increased alanine and lowered progesterone and (3) both the MF0048 pathway (involved in serine degradation) and MF0049 pathway (involved in threonine degradation) increased glutamic acid (see GitHub for relevant <u>figures</u> and <u>results</u>). However, there were high levels of heterogeneity across almost all analyses and the exposure and outcome data were obtained from GWAS analyses in the same individuals (see also “*Quality assessment*” below).

The Liu *et al*.^25^ study, focusing on optic neuritis, undertook discovery and replication analyses with 197 unique gut microbial traits using one or both of two exposure data sources (i.e., from Goodrich *et al*.^34^ and Kurilshikov *et al.*^10^), where 47 of these gut microbial traits were analysed using both exposure data sources. Summary statistics for the outcome were sourced from GWAS analyses conducted in either FinnGen (https://r7.finngen.fi/) or UK Biobank^35^. However, when the exposure data were sourced from the GWAS conducted by

Goodrich *et al*.^34^, only the FinnGen data were used; therefore, only the analyses using the MiBioGen data as the source of the SNP-exposure associations (and therefore both outcome data sources) were meta-analysed and presented here (see GitHub for relevant <u>figures</u> and <u>results</u>), representing 196 unique gut microbial traits. Results provided some evidence that higher abundances of bacteria within the *Eubacterium rectale* group, *Hungatella* genus and the UCG003 group in the *Ruminococcaceae* family reduced the risk of optic neuritis and bacteria within the UCG010 group in the *Lachnospiraceae* family increased the risk of optic neuritis. However, neither would have reached a multiple testing corrected p-value given the number of gut microbial traits being analysed in this study (see also “*Quality assessment*” below).

Finally, the study by Xu *et al*.^27^, assessed the causal role of gut microbial traits on several autoimmune diseases including systemic lupus erythematosus (SLE), rheumatoid arthritis (RA), inflammatory bowel disease (IBD), multiple sclerosis (MS), celiac disease (CeD) and type 1 diabetes (T1D) using the MiBioGen consortium as the source of information for the SNP-exposure association and data from outcome-specific GWASs in UK Biobank. Full MR results were reported only for the discovery analyses, which relied on previously published GWAS datasets as outcome sources (i.e., one GWAS per outcome). Replication analyses were also only presented for two gut microbial traits (bacteria within the *Bifidobacterium* and *Ruminococcus* genera) and all outcomes except RA (as authors stated that there was “no evidence” for a causal effect of microbial traits on this outcome in the discovery sample). However, data used for replication analyses for CeD (Dubois *et al*.^36^) and SLE (Julià *et al*.^37^) were not independent from those used in the discovery analyses (i.e., there was substantial sample overlap in the GWASs). Therefore, we did not consider these to be replications. Of those findings that were replicated, meta-analysis of both discovery and replication analyses provided little evidence for an effect of bacteria in either *Bifidobacterium* or *Ruminococcus* genera on IBD, MS or T1D (see GitHub for relevant <u>figures</u> and <u>results</u>), where there were high levels of heterogeneity across all analyses (see also “*Quality assessment*” below).

##### Between-study replication

When comparing studies, there were 5,768 estimates of the 48,082 total across the 66 studies that represented analyses of the same exposures (284 unique exposures) and same outcomes (193 unique outcomes). Of these, nearly all (94.9%; n=5,475) had unclear or unspecified exposure units, and 9.7% of estimates (n=558) lacked clear outcome units. As a result, 98.2% of these estimates (n=5,666) could not be meta-analysed due to insufficient clarity on how the MR estimates should be interpreted. For example, despite many studies stating that the MR estimate reflected a difference in the “abundance” of microbial traits, they did not specify the exact unit (e.g., SD or taxonomical counts) and many studies did not report the exposure measure or unit at all (see “*Quality assessment*” below).

Of those 102 estimates that had comparable exposures (and units) and outcomes (and units), only nine estimates (i.e., 0.2% of the 5,768 estimates) used different outcome study sources across studies, making them eligible for meta-analysis. These nine estimates were from two studies (Hughes *et al*.^9^ and Ning *et al*.^38^) related to the “Brain” outcome group and specifically used two-sample MR analyses to estimate the causal effect of higher abundances (in SD units) of bacteria within the *Bifidobacterium*, *Butyricicoccus* and *Parabacteroides* genera (sourced from both the GWASs published by Hughes *et al*.^9^ and Kurilshikov *et al*.^10^) on the odds of either/both Alzheimer’s disease (sourced through GWASs published by Lambert *et al*.^39^ and Jansen *et al*.^40^) and Parkinson’s disease (sourced through GWASs published by Simón-Sánchez *et al*.^41^ and Nalls *et al*.^42^).

The MR analyses conducted by Hughes *et al*.^9^ used a single instrument selection strategy, using the SNPs that past a genome-wide p-value threshold (p<2.5×10^−08^) in the GWAS conducted in the same study, whilst Ning *et al.*^38^ conducted analyses comparing two instruments selected based on a genome-wide p-value threshold (p<5×10^−08^) and a lenient p-value threshold that has become commonplace in the application of MR to gut microbiome exposures (p<1×10^−05^; see Box 2 and “*Quality assessment*” below). Using the lenient p-value threshold in Ning *et al*.^38^ and stringent p-value threshold in Hughes *et al*.^9^, the MR estimates were directionally consistent, where higher abundances of bacteria in the *Butyricicoccus* genus reduced the risk of Alzheimer’s disease and Parkinson’s disease, and higher abundances of bacteria in the *Parabacteroides* genus increased the risk of Parkinson’s disease. However, meta-analysis provided little evidence for a strong causal effect in any relationship (see GitHub for relevant <u>figures</u> and <u>results</u>).

When comparing the results using the more stringent p-value threshold (p<5×10^−8^) used by Ning *et al.*^38^ with the analyses undertaken by Hughes *et al*.^9^, the analyses of bacteria in the *Bifidobacterium* genus and Alzheimer’s disease indicated that a higher abundance of this microbial trait increased Alzheimer’s disease by 0.6% (meta-analysed odds ratio (OR) from random effects model: 1.06; 95% CI: 1.02, 1.09; I-squared and p-value for heterogeneity: 0% and 0.45, respectively). However, it is worth noting that, whilst the GWASs from which the Alzheimer’s disease data were sourced were different, there is a substantial overlap in the cohorts that contributed to those GWASs (e.g., with both GWASs comprising data from the International Genomics of Alzheimer’s Project (IGAP) including the European Alzheimer’s disease Initiative (EADI), Alzheimer Disease Genetics Consortium (ADGC), Cohorts for Heart and Aging Research in Genomic Epidemiology consortium (CHARGE) and Genetic and Environmental Risk in AD consortium (GERAD) data). Therefore, these analyses are not truly independent and the overlap in outcome study samples may indeed explain the consistency in effect estimates between these studies.

#### Quality assessment

##### Across all studies

Using our bespoke 19-item quality assessment tool, the mean number of items that were judged as “low quality” across all studies was 6.12 (median = 6.00; SD = 1.89; range = 2-10). The mean number of items that were judged as “moderate quality” across all studies was 10.68 (median = 11.00; SD = 2.11; range = 5-16) and the average number of items that were judged as “high quality” across all studies was 2.20 (median = 2.00; SD = 2.03; range = 0-12). In all studies, at least one item was judged as “low quality”; therefore, the overall quality of all studies was considered low (Figure 4, Supplementary File 3).

**Figure 4.**
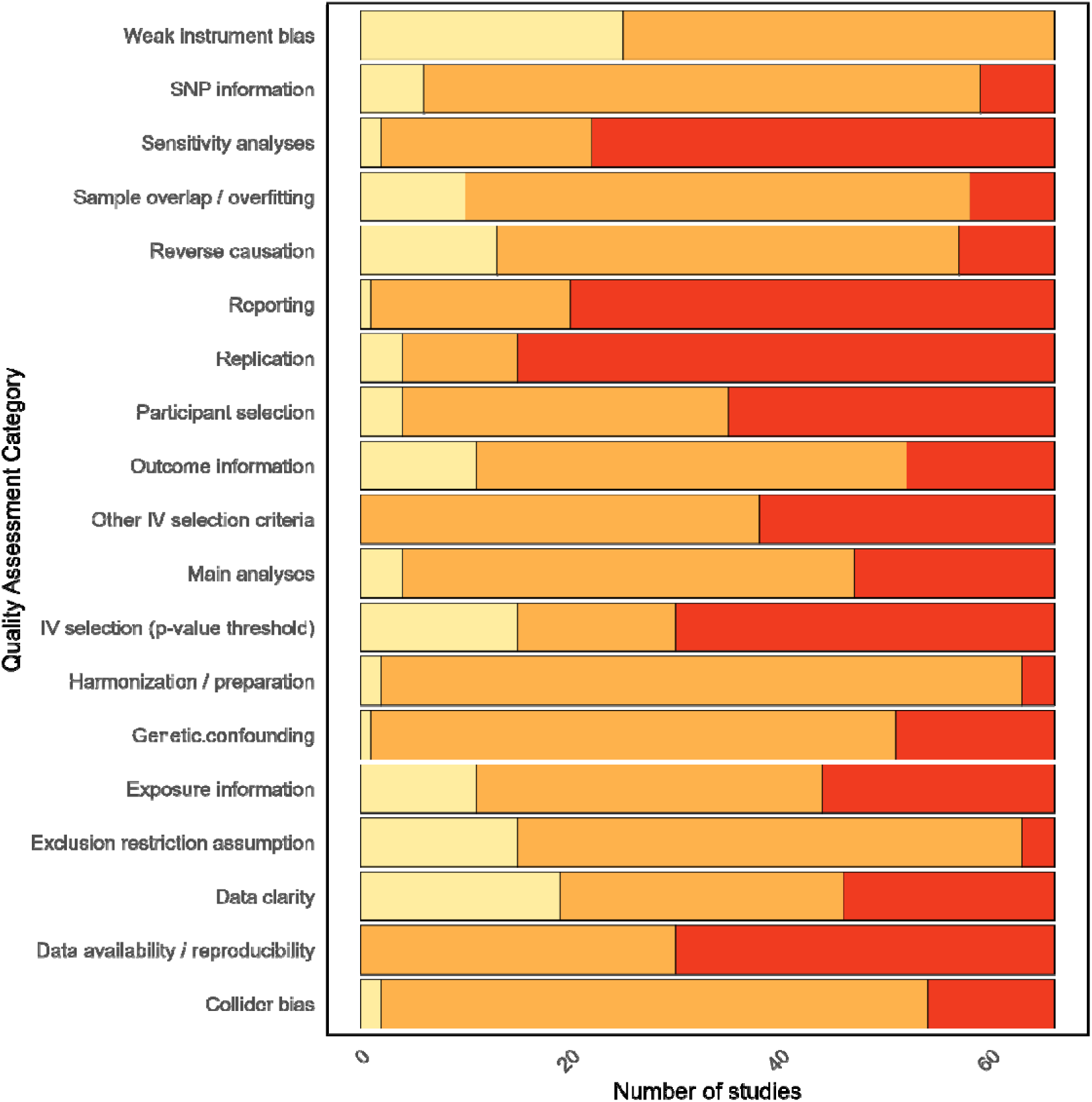
Quality assessment results across all studies. Number of studies (x-axis) that were considered “high quality” (yellow), “moderate quality” (orange) or “low quality” (red) across items on the quality assessment tool (y-axis). Low quality indicated where a study undertook an analysis correctly or made a correct methodological decision, moderate quality indicated where an item on the quality assessment tool was unclear within a study or a study undertook an analysis or made a decision that was likely to lead to a minimal level of bias, and high quality indicated where a study undertook an analysis explicitly incorrectly or made an unjustified and incorrect decision.

The items in the quality assessment tool on which studies were most likely to be judged as “low quality” were those relating to: replication (with 77.3% of studies being judged as “low quality” on this item; Box 2); reporting (69.7% of studies); sensitivity analyses (66.7%); p-value threshold for IV selection (54.5%); reproducibility (54.5%), selection of participants (47%) and other IV selection criteria (42.4%). Where studies were judged as “moderate quality” based on one of the items in the quality assessment tool, there was usually either uncertainty when considering the specific item or there was a mixture of approaches used that were partially appropriate (i.e., not incorrect enough to be considered “low quality” and not certain enough to be considered “high quality”). The items on which most studies were considered “moderate quality” related to harmonization and preparation of data for analyses (with 92.4% of studies being judged as “moderate quality” on this item), information about the number of SNPs used in analyses (80.3% of studies), bias either due to population-level confounding or collider bias (78.8%), genetic confounding (75.8%), testing of the exclusion restriction criteria (72.7%), overfitting due to sample overlap (72.7%), reverse causation (66.7%), reporting of main analyses (65.2%), outcome information (62.1%), weak instrument bias (62.1%), other IV selection criteria (57.6%), exposure information (50%), selection of participants (47%), and data availability and reproducibility (45.5%).

Very few studies were of “high quality” in many of the items of the quality assessment tool (Figure 4). The item on which the largest number of studies were considered “high quality” related to weak instrument bias (37.9% of studies), data sources (28.8%), p-value threshold for IV selection (22.7%), assessment of the exclusion restriction criteria (22.7%) and reverse causation (19.7%). See Box 2 for common reasons for a study being judged as “low”, “moderate” or “high” quality across items of the quality assessment tool and justifications for this judgement. Full details of the quality assessment of each study across all 19 items can be found in Supplementary File 3.

##### Within studies

When focusing on individual studies, there were five where approximately half (i.e., between 47.4-52.6%) of the items on the quality assessment tool were considered “low quality”^10,43–46^, and an additional 12 studies where 42.1% of the items were considered “low quality” (Figure 5)^31,47–57^. Of these, nine were also included in the set of 14 studies where none of the items were considered “high quality”^44,47–49,51,52,54–56,58–62^. However, over half of these 14 studies^44,49,51,55,56,58,59,61^ were conference abstracts or correspondences; therefore, many details necessary for a full quality assessment were unclear given the small number of words permitted compared to full texts.

**Figure 5.**
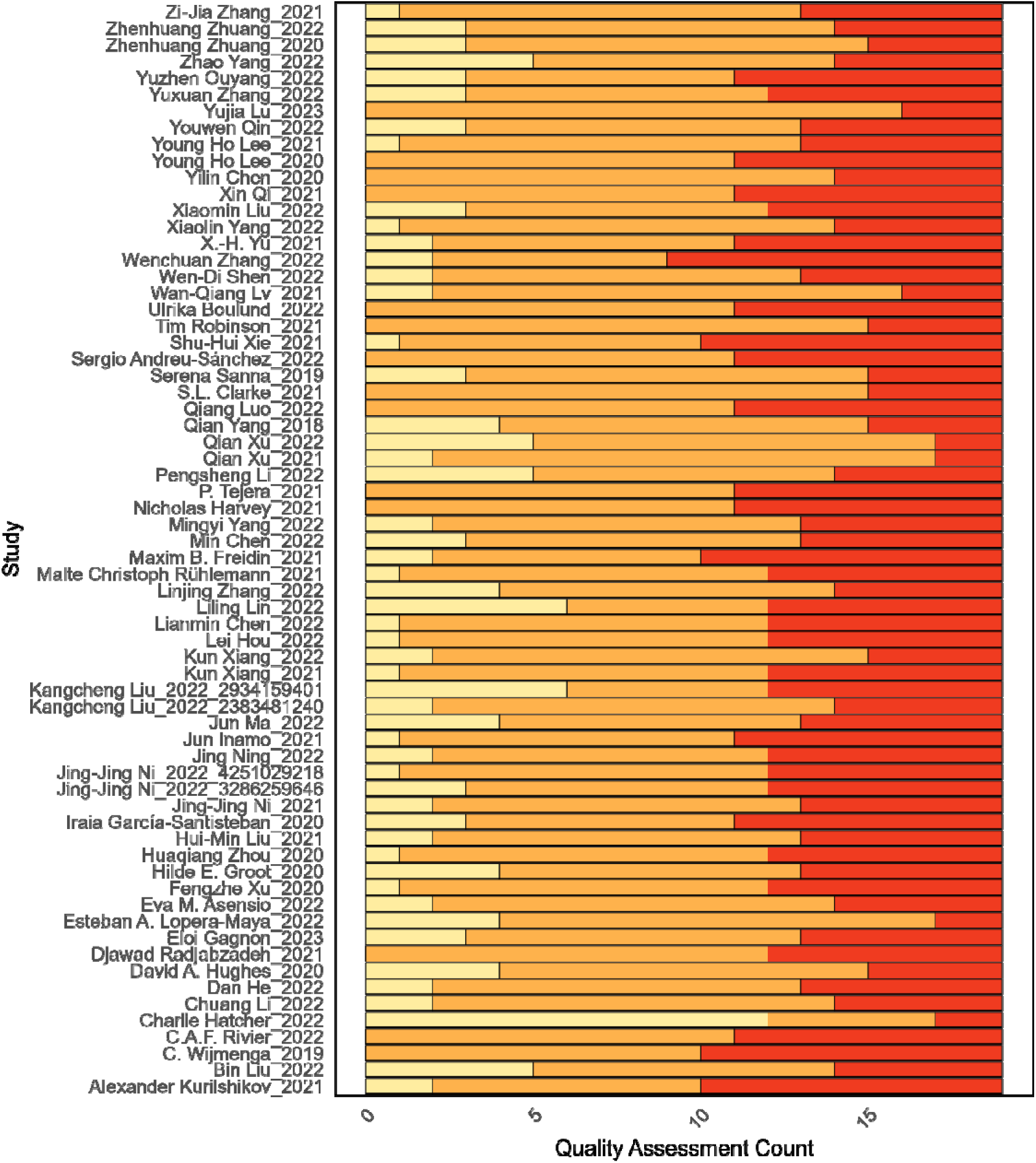
Quality assessment results within each study. Number of items on the quality assessment tool (x-axis) on which each study (y-axis) was considered “high quality” (yellow), “moderate quality” (orange) or “low quality” (red). Low quality indicated where a study undertook an analysis correctly or made a correct methodological decision, moderate quality indicated where an item on the quality assessment tool was unclear within a study or a study undertook an analysis or made a decision that was likely to lead to a minimal level of bias, and high quality indicated where a study undertook an analysis explicitly incorrectly or made an unjustified and incorrect decision. Note that there were two publications by two of the same authors (i.e., Liu 2022 and Ni 2022); therefore, to separate each pair clearly, the study identifier was added as a suffix to each study accordingly.

Below is a description of the most common weaknesses across all studies that led to them being judged as “low quality”, with reference to the items on the quality assessment tool (also see Box 2 for more details and Supplementary File 3 for a comprehensive overview of the results of the quality assessment across all studies). Within Box 3, we have collated a series of recommendations for best practice when undertaking MR analyses assessing the downstream causal relevance of gut microbial phenotypes.

#### Instrument selection and weak instrument bias

At least 36 studies (54.5%) included in this review used a lenient p-value threshold (i.e., a p-value larger than the widely accepted genome-wide significant threshold of p < 5×10^−08^) to select SNPs associated with gut microbial traits. Many of these studies referenced Sanna *et al*.^24^ (the first published study to take such approach) as justification^10,23,24,26,28–31,45–48,52–54,57,60,62–80^. Whilst this has become commonplace in the application of MR with the gut microbiome, it is not best practice as it leads to explicit violation of the first MR assumption and likely induces bias via violations of further MR assumptions. This is particularly problematic in the context of the high level of heterogeneity observed across mGWASs and their signals, where there is very limited overlap^81^. One study of note involved identifying the p-value threshold at which the power of independent SNPs to predict the microbial trait of interest was greatest (i.e., by maximising the r-squared describing phenotypic variance explained)^46^. However, the use of such metrics does not guarantee that the SNPs will be valid for MR analyses. Indeed, whilst increasing the number of SNPs in an MR instrument tends to increase the r-squared value, inclusion of more SNPs increases the likelihood of inducing type I error and bias via horizontal pleiotropy, thereby reducing the validity of the instrument selected.

In addition to using a lenient p-value threshold, many studies used further unjustified and inappropriate instrument selection criteria. Firstly, at least 15 studies^10,23,25,27,38,45,48,52,53,65,70,75,79,82,83^ removed SNPs that had an F-statistic <10. Despite this ensuring that instruments are “strong” (and indeed, no studies included in this review had weak instruments), this is not best practice - the F-statistic should be used as a marker of bias when interpreting results rather than instrument selection criterion (Box 2, Box 3)^84^. Secondly, at least 14 studies removed palindromic SNPs^25,27,29,45,53,64,65,68,70,75,80,85–87^, possibly needlessly given that the effect allele frequencies were seemingly available, a decision that likely lowered power in analyses given the already small number of SNPs available. Thirdly, at least 22 studies used clumping thresholds that were likely too lenient for MR analyses^24,26–31,45,47,53,54,60,67,68,70,72–74,77,79,88,89^, which will have impacted the interpretation of estimates given that the methods used did not account for correlation amongst SNPs. Additionally, there were two further studies that, whilst using a genome-wide p-value threshold (i.e., 5×10^−08^) to select SNPs associated with the gut microbiome (which is best practice), the SNPs used were clearly not independent (as seen rather strikingly in their scatter plots), which will have certainly led to biased estimates and standard errors.

Lastly, at least 11 studies undertook pleiotropy-robust and outlier tests before performing the main analyses^27–29,45,67,68,70,75,82,90,91^, which should ideally be performed as sensitivity analyses testing the robustness of main analyses rather than for identifying SNPs for exclusion beforehand. Related to this, at least 12 studies^25,28,38,52,63,64,67,73,77,86,89,92^ excluded SNPs before analyses due to being associated with either the outcome of interest or phenotypes that could be confounders of the exposure and outcome, with the aim of complying with the second MR assumption. This suggests a misunderstanding of the second MR assumption (Figure 1) and the exclusion of SNPs based on such associations could very likely be unnecessary and could lead to an underestimate of the causal effect if, for example, such associations reflect vertical pleiotropy (i.e., the very causal pathway by which the microbiome influences the outcome).

#### Genetic confounding

Fifty of the studies included in this review (75.8%) did not discuss the impact of population-level or genetic confounding on MR estimates (i.e., the second MR assumption)^10,22–24,26,27,29–31,43–51,53–62,65,66,69–74,76,78–80,82,85,87,88,90,91,93–96^. Whilst a further 15 studies explored individual-level confounding factors (e.g., assessing the associations between SNPs and confounders of the exposure-outcome association or undertaking multivariable MR adjusting for correlated phenotypes, as described above), this related somewhat to the misspecification of the second MR assumption^9,25,26,28,29,38,52,63,64,67,68,75,77,83,86,87,89,92^. Whilst such individual-level confounders can be used as indicators of population-level confounding factors, this was seemingly not the intention for many of these studies. Indeed, such analyses relate more to possible bias due to horizontal pleiotropy through confounders of the exposure-outcome association (referred to as heritable confounders or correlated pleiotropy)^1,97^. Only one study included in the review undertook colocalization analyses to understand whether the causal effect estimates were biased due to genetic confounding (i.e., where the exposure-related SNP was in LD with an outcome-related SNP)^98^.

#### Collider bias

At least 12 studies either used publicly available GWAS analysis results or performed their own GWAS analyses that were adjusted for heritable covariates^22–24,26,30,46,48,64,72,75,82,90^. Of these, at least one MR study using two-sample methodology used summary statistics from GWASs of the exposure and outcome, where analysis adjustments were substantially different between data sources^72^. These processes can introduce bias in MR effect estimates due to including variables that may lie on the causal pathway (thus, possibly inducing collider bias and/or confounding) and inconsistencies in the structure of data contributing to MR effect estimation^99^.

#### Assessment of the exclusion restriction assumption

Three studies did not undertake any sensitivity analyses to test for the robustness of their MR estimates to violations of the third MR assumption (i.e., no horizontal pleiotropy)^44,46,49^. However, it is worth noting that two of these were conference abstracts with very little space for detail^44,49^. Of the 66 studies included in this review, 15 conducted sensitivity analyses that were well justified and clearly described within the methods section of the paper with regards to their assumptions and application^24,25,63,66,68,71,75–77,79,83,85,89,92,98^. The rest of the studies (n=48) had performed such analyses, but these were not transparently justified or reported, with many using methods typically used as sensitivity analyses in conjunction with main analyses without comparative discussion around their different assumptions^9,10,22,23,26–31,38,43,45,47,48,50–62,64,65,67,69,70,72–74,78,80,82,86–88,90,91,93–96^.

#### Population structure, sample overlap and overfitting

Thirty-one of the studies included did not provide transparent information regarding the population structure of both the exposure and outcome data sources (and particularly, their similarity) and whether or how such population structure had been handled in the respective GWASs (e.g., through inclusion of principal components)^9,28–31,43,44,51,54–56,58,59,61–64,66,67,73,74,82,83,86,88,90,91,93,95,96,98^. Information provided in 31 other studies indicated that the population structure across exposure and outcome datasets were heterogeneous, which may lead to a violation of the second MR assumption^10,23–27,38,45,47–50,52,53,57,60,65,68–70,75–77,79,80,85,87,89,92,94^. For example, many studies used the MiBioGen consortium, which is a GWAS of the gut microbiome in individuals of mixed (but predominantly European) ancestry, as the source of instruments for the microbiome and data from individuals of mostly European or sometimes mixed or even totally different ancestries as the source of outcome information, without commenting on the possible bias that this may induce due to differential population structure across data sources.

Conversely, at least six studies included analyses whereby they conducted a GWAS of both the exposure and outcome using the exact same participant data that was then used in MR analyses (and, in the case of one-sample MR analyses, the weights from the exposure GWAS were then used in the MR analyses)^22,26,30,46,72,78^. There were also two additional studies that used data for two-sample MR analyses where participants from exposure and outcome GWASs explicitly overlapped, but the bias induced was not assessed^47,79^. Therefore, whilst the population structure amongst the data used for the GWAS and MR analyses is homogenous in these cases, the overlap in participants will likely bias MR estimates (and their precision) due to overfitting. Additionally, estimates derived specifically from two-sample MR analyses will also be biased towards the observational analysis with weak instrument bias (and, in many cases, tests for instrument strength were not provided). Only ten studies explicitly tested for and described a lack of sample overlap or problems with overfitting^25,53,64,75–77,83,89,96,98^; however, many studies (n=48) did not provide enough information to discern whether population overlap and overfitting was an issue^9,10,23,24,27–29,31,38,43–45,48–52,54–63,65–71,73,74,80,82,85–88,90–95,100^.

#### Reverse causation

Thirty-eight of the studies included in this review did not provide any description of tests for potential bias driven by reverse causation, whereby the observed effect in an MR analysis may be explained by an effect of the IV on the outcome which then, in turn, influences the gut microbial exposure (i.e., the IV is invalid)^22,23,25,28–31,43,44,47–52,54–56,58,61–67,69,74,77,79,82,85,87,89–94^. Whilst thirteen studies tested for reverse causation, typically via undertaking a formal MR analysis in the reverse direction, and presented these results^27,46,53,68–71,75,76,80,86,95,98^, an additional six studies undertook such analyses but did not present the results clearly (or at all) in the text^9,10,24,59,72,83^. Nine studies undertook analyses testing for bidirectionality or reverse causation; however, the results presented were incorrectly and misleadingly interpreted. For example, many studies stated that there was evidence for an effect between a particular phenotype and a gut microbial trait, but only presented p-values instead of reflecting on the directional consistency of beta-coefficients derived across different MR methods (where, in some cases, the methods were directionally inconsistent suggesting a lack of robust evidence for an effect)^26,45,57,60,73,74,78,88,96^. Additionally, whilst one study indicated that they performed a reverse MR analysis, suggesting no evidence for reverse causality, their results described that they found no “common SNPs” between cardiovascular proteins and bacterial features, which is not a method for identifying reverse causality (given differential power in GWAS data sources)^45^. Plus, the results were not presented.

#### Harmonization process

Almost all of the studies (92.4%) failed to provide clear information about the pre-analysis harmonization process in their MR analyses. This is an established and crucial step for ensuring MR estimates are correct, ensuring that changes in the exposure and outcome variables are aligned to the same effect allele^101^. Whilst only two studies provided sufficient information about harmonization^31,82^, the results of three studies indicated that harmonization may have been undertaken incorrectly^10,57,72^. Specifically, the x-axes of scatter plots presented included negative values, which is not necessarily an issue for the analyses depicted in those plots (i.e., the inverse variance weighted and weighted median approaches, where the intercept is restricted to zero) but, given that authors also undertake and present estimates using the MR-Egger method (which requires all SNP-exposure estimates to be directionally consistent), it is unclear whether the data were harmonized appropriately.

#### Presentation of main and sensitivity analysis methodologies

In terms of the presentation of the results, many studies failed to provide fully interpretable effect estimates (e.g., with clear units) from the main analyses. Specifically, 19 studies either did not report any results for main analyses or only presented p-values for these results, which is not informative for understanding the direct and likely magnitude of the causal effect estimate^22,25,31,43–45,47,50,53–55,57,72,74,77–79,82,83^. Equally, at least 44 studies failed to either apply or correctly and comprehensively interpret sensitivity analyses^9,10,22,23,25,28,29,31,38,43–49,51–53,55–58,60,63–65,67,69,72–76,78,82,85–88,90,93,94,96^. If sensitivity analyses were applied, studies either incorrectly and misleadingly stating that there was no evidence of bias or that results across sensitivity analyses corroborated the main findings, a decision that was informed by p-values alone, despite many inconsistencies across effect estimates from different pleiotropy-robust methodologies. For example, many of these studies stated that there was no evidence of bias due to heterogeneity or pleiotropy (e.g., by interpreting the Cochran’s Q statistic, MR-PRESSO test and MR-Egger intercept, where p-values were >0.05); however, effect estimates across different methodologies were inconsistent with main effect estimates, which would question the validity of main effects. Similarly, the interpretation of leave-one-out analyses was also incorrect in at least nine studies^25,64,67,72,75,76,82,85,87^. Specifically, these studies concluded that no single SNP influenced the initial results, despite the removal of at least one SNP attenuating the main effect, suggesting that the initial effect estimate may have indeed been driven by those SNPs.

#### Replication

Whilst four studies undertook formal replication analyses (see *Meta-analysis* above)^24–27^, most studies either did not undertake a formal replication analysis at all or attempted such an analysis but, due to a lack of independent data sources, it could not be considered a formal replication (n=51)^9,10,23,28,31,38,43,45,48–59,61–63,65–71,73–77,79,80,83,85–96,98^. Additionally, 11 studies coupled MR analyses with another study design assessing the same association (e.g., a case-control or cross-sectional study)^29,30,44,46,47,60,64,72,78,82,91^, some of which did not directly compare results across analyses. However, whilst useful comparative analyses, these cannot be considered formal replications of the initial MR analyses undertaken.

#### Reporting of data

Whilst information provided about the data used for both the exposure and outcome was clear and matched external sources in 19 of the 66 studies^22,27,28,30,31,45,53,63,65,67,75–78,82,86–88,94^, this was not the case for the remaining studies, where there were inconsistencies and uncertainties about what exact data were used for MR analyses. This was the case for either one data source (e.g., either the exposure or outcome data source for a two-sample MR analysis) for 27 studies^9,10,23–27,29,38,47,48,52,57,64,69–73,79,80,85,90–92,95,98^ or for all data sources for 20 studies^43,44,46,49–51,54–56,58–61,66,68,74,83,89,93,96^. Many studies did not provide clear details about the measurement and units of the exposure and outcome, information that is required for interpretable MR-derived effect estimates. Specifically, 22 studies failed to provide adequate information about the measurement and unit of interpretation for the exposure^29,31,43–45,47,50–56,63,66,67,71,85,89,93,94,96^ and 14 studies similarly failed to provide the same information about the outcome^10,29,31,43–45,48–51,55,56,60,73^. Nine of those studies failed to provide information on both the exposure and outcome^29,31,43–45,50,51,55,56^.

Most studies provided information on the measurement of the gut microbiome but did not specify whether the features analysed were reflective of, for example, the abundance or presence vs. absence of gut microbial traits, and did not provide information on the specific unit, whilst others provided information about the measurement but not the unit. Similarly, most studies provided information about the measurement and definition of the outcome but not the unit to allow transparent interpretation of the MR estimates. For example, estimates presented in one study were unclear, where ORs are presented for the effect of the gut microbial traits on continuous outcome variables^43^. Similarly, the units were particularly unclear in two studies, where results were presented for the effect of the “gut microbiome” or “functional units” on an outcome of interest^58,89^. Additionally, no studies commented on the likely complexities of analysing composite phenotypes (i.e., when using relative abundances), whereby the distribution and thus unit may not be equivalent across GWASs and, thus, may not be directly comparable^102^.

Information about SNPs used in analyses was often provided but, for 53 studies^10,23–25,27,30,38,43,45–55,57,58,60,62,63,65–72,74–80,82,83,85–96^, it was unclear as to how the exact number used was attained and, for seven studies, information on the number of SNPs used in analyses was not provided at all^22,26,44,56,59,61,73^. In contrast, six studies made this very clear^9,28,29,31,64,98^, where some used flow-chart-like figures describing how many SNPs were removed and at what stage of the pre-analysis pipeline.

#### Reproducibility and comparison of studies undertaking the same analyses

With regards to overall reporting, 46 studies did not transparently or accurately present effect estimates, which in turn meant that conclusions were often misleading^10,22–26,28,29,31,38,43–49,51–58,60,63–65,67,69,70,72–74,76,78,82,85–88,90,93,94,96^. An additional 19 studies reported effect estimates more transparently, where, sometimes, not all results presented, but the interpretation of the results (e.g., in terms of unit) could have been clearer^9,27,30,50,59,61,62,66,68,71,75,77,79,80,83,89,91,92,95^. Approximately half of the studies did not provide clear information (or direct links to) data sources or code used for analyses^9,10,23,25,38,43,44,46–52,54–56,58,59,61,62,64,66,68,69,71,80,83,85,89–93,95,98^, whereas the other half of studies either provided reproducible information about the data sources used for the analyses or links to the analytical code^22,24,26–31,45,53,57,60,63,65,67,70,72–79,82,86–88,94,96^. Overall, neither the methods nor results were reported transparently or reproducibly for any study.

To highlight the importance of transparent and reproducible reporting and test whether the application of MR was comparable amongst this selection of studies, we searched for and compared results where reporting suggested ostensibly the same analyses had been performed. Of the 5,768 estimates across the 66 studies that represented analyses of the same exposures (284 unique exposures) and same outcomes (193 unique outcomes), 4,085 estimates were conducted using the same datasets. However, of these, only four estimates derived from Xu *et al*.^27^ and Xiang *et al*.^85^ were directionally comparable and reflected the analysis of two exposures (specifically, bacteria within the *Allisonella* and *Bifidobacterium* genera) and one outcome (specifically, systemic lupus erythematosus, SLE). The other 4,081 estimates were either derived using unclear data sources (i.e., appeared as “NA”), did not have comparable units, or used different strategies and methodologies (e.g., a different p-value threshold for instrument selection); thus, were not reflective of the same analysis.

Firstly, it is worth noting that neither study clarified the specific exposure unit but, considering they used summary-level data for the exposure from the same mGWAS, it is highly likely that they reflect the same unit change in each of the two bacterial traits (i.e., standard deviations). Similarly, whilst Xu *et al*.^27^ provided information about the outcome measurement, beta-coefficients (rather than ORs) were provided and it was not made clear whether these were logORs and, conversely, whilst Xiang *et al*.^85^ did not provide information on the outcome measurement, effect estimates were presented as ORs.

The derived estimates assessing the effect of bacteria within the *Allisonella* genus on SLE using one SNP (rs602075) were consistent across both studies (Supplementary Figure 2 and see GitHub for relevant <u>results</u>), where slight differences were due to one study presenting results as rounded ORs and the other as rounded logORs. Whilst these estimates are consistent, there were inconsistencies in some of the information provided across studies, which questions the total transparency, reproducibility and clarity of methods and results. For example, both studies provided the SNP-exposure and SNP-outcome summary-level data for SNPs used in analyses in their supplementary tables. However, there were differences in the p-values for the SNP used in MR analyses for *Allisonella* (i.e., with Xu *et al*.^27^ stating that the p-value of the SNP-exposure association was 3.57×10^−08^ and Xiang *et al*.^85^ stating that it was 1.77×10^−08^).

Whilst the derived estimates assessing the effect of bacteria within the *Bifidobacterium* genus on SLE were directionally consistent with a protective effect, there was a larger difference between estimates compared to that with *Allisonella* (Supplementary Figure 2). Authors used the same data for the exposure and outcomes, conducted the same analyses and used the same p-value threshold for instrument selection (i.e., 5×10^−08^); however, the reasons for this difference between estimates are likely due to differences in other instrument selection criteria. Xiang *et al*.^85^ estimated the causal effect of bacteria within the *Bifidobacterium* genus using two SNPs (rs182549 and rs7322849) that were selected based on having a minor allele frequency (MAF) >0.01 and an F-statistic >10 that were clumped to retain independent SNPs (using an r-squared <0.001 and genomic distance of 10,000kb). Additionally, palindromic SNPs were removed. Similarly, Xu *et al*.^27^ also removed palindromic SNPs and restricted the set of SNPs to those that had an F-statistic >10; however, they did not mention any restrictions based on MAF and used a different and much more lenient clumping threshold (using an r-squared <0.01 and genomic distance of 500kb). This resulted in the use of six SNPs (rs182549, rs4954567, rs6723108, rs6759321, rs7322849, rs7570971), two of which were the same SNPs as used in Xiang *et al*.^85^.

Despite using two of the same SNPs across studies, there were also discrepancies in the summary-level data provided for the *Bifidobacterium*-related SNPs as for the *Allisonella*-related SNP. Firstly, the effect allele and other allele for rs7322849 were flipped (i.e., in Xu *et al*.^27^, the effect/other alleles were specified as being T/C, respectively, and, in Xiang *et al*.^85^, these were specified as being C/T, respectively) and, whilst the SNP-exposure effect estimate was the same in the supplementary table, it is unclear whether this may have generated opposing MR estimates across studies if, indeed, the effect alleles used in analyses were incorrectly specified. Secondly, the p-values for the SNP-exposure associations were also different, despite authors reporting that these were obtained from the same data. Lastly, given that Xu *et al*.^27^ used more (and likely weaker and invalid) SNPs in MR analyses, the effect estimate derived was stronger than that derived in Xiang *et al*.^85^ using two SNPs, as weak instruments are more likely to bias effect estimates away from the null (more specifically, towards the observational effect estimate, which in turn is more likely to be biased by confounding; Box 1). Therefore, despite sharing the same data and approach, the inconsistency between these two estimates reiterates the importance of reproducibility, transparency and, specifically, choice of instruments and selection criteria for undertaking MR analyses, where making data and code available is ideal.

Despite this, there were several studies that were either infrequently scored as “low quality” or most frequently scored as “high quality” across the quality assessment tool (Hatcher *et al*.^98^, Lopera-Maya *et al*.^95^ and Xu *et al*.^27^). Whilst none of these studies are totally reproducible or flawless, they demonstrate good standards that future studies should emulate in efforts to improve the appropriate application and quality of reporting MR studies assessing the causal role of the gut microbiome on human health outcomes. These best practices are summarised in Box 3.

## DISCUSSION

In summary, according to our quality assessment tool, all studies included in this review were of low quality, due to the inappropriate application of MR – specifically, instrument selection, exposure and outcome definition, choice of analytical methodology, assessment of reverse causation and replication – and lack of transparent reporting of findings (see Box 3). In this systematic review, we aimed to investigate how MR has been applied to assess the causal relevance of the gut microbiome in human health and evaluate the quality of these studies. We identified 66 studies, which comprised 48,082 individual MR estimates of the relationship between 612 gut microbial traits (defined by relative abundance, presence vs. absence or functional pathway) and 905 health outcomes including those categorized into autoimmunity, behaviour, cancer, prescription drug usage, immunity, inflammation, longevity, medical procedures, metabolic health, nutrition, pain, sexual and reproductive health, and diseases of several organs and systems. The consistently poor quality of studies, along with the limited caution in interpreting estimates and widespread disregard of core MR assumptions, suggests that many of these studies, some published in reputable journals, should not have been brought forward for publication and reflects poorly on a field that attempts to maintain high standards. Therefore, given the overall quality, the results presented here, especially those from within-study and between-study meta-analyses, should be interpreted with extreme caution.

The overall low quality of studies included in this review is particularly alarming given the growth in the application of MR to understand the causal role of the microbiome in human health. As recently summarised by Cupido *et al*.^103^, the growth in such studies being published is exceptionally fast – i.e., they mentioned that, in 2022, there were 43 papers in PubMed using the terms “mendelian randomization” and “microbiome”, a number which increased to 267 in 2023 and over 353 in July of 2024. As of June 2025, the same broad search strategy astoundingly produces over 860 studies. This, coupled with the overall low quality of papers where misinformation may lead to inappropriate conclusions and recommendations, is particularly alarming given the discrepancy in the size of this body of work and the level of clarity (or lack thereof) in the preventative and treatment utility of the gut microbiome. Thus, identifying high quality studies will become even more challenging in the future due to the increasingly high volume of potentially inaccurate and misleading studies.

Whilst this current review only formally captures studies published up to January 2023, the quality of the literature has, in our opinion, not noticeably improved and the conclusions of this review (the primary aim of which was to review, and assess the quality of, the existing evidence, and not to summarise the causal effect of the microbiome on health outcomes) would not change. Given the rate at which MR studies on this topic are being published, it would also be impractical to update a review of this kind in a timely manner. Nevertheless, the results presented here represent a snapshot of the literature on how MR is being applied to assess the downstream effect of the microbiome. There are several other limitations to this work. Firstly, based on our criteria for meta-analysis, estimates were excluded if there was any overlap between the outcome data sources between studies; however, it is possible that this was not completely accurate given that not all studies comprehensively reported the studies used in their analyses or estimated participant overlap between studies used. Secondly, given the sheer number of estimates across the 66 included studies and the fact that results tended to be buried in supplementary materials and not described transparently, and despite best efforts, manual data entry may have introduced unintentional inaccuracies. Thirdly, this review focused on measures of the gut microbiome specifically (i.e., bacterial traits) and did not include any microbiome-related or derived metabolites explicitly. However, the conclusions presented here regarding caution in the application and interpretation of MR analyses is still very much applicable to such traits, as with all complex -omics phenotypes (as highlighted similarly by Cupido *et al*.^103^), where there is often high-dimensional data with few related SNPs most of which are of unknown biological relevance.

In conclusion, whilst MR is a useful method that, when applied appropriately and compared with findings from other study designs, can guide causal inference, most of the current studies applying MR to understand the relationships between the gut microbiome and health outcomes are of poor quality and potentially highly biased. Our systematic review highlights the requirement of a basic understanding of MR methodology; more comprehensive sensitivity analyses testing the validity of findings; more caution in the interpretation with more replication; and triangulation efforts with varying study designs, given the complexity of host genetic effects on the gut microbiome. These requirements, alongside rigorous editorial processes to check that the work adheres to best practice in MR analyses, are necessary to truly understand the causal relevance of the microbiome in human health and disease.

## Data Availability

The data produced as part of this review are presented in the paper supplement and online at https://github.com/KaitlinHazelWade/Microbiome_MR_review/.

## Ethics approval

Ethical approval was not required for this study.

## Transparency statement

As authors affirm that this manuscript is an honest, accurate, and transparent account of the study being reported; that no important aspects of the study have been omitted; and that any discrepancies from the study as planned (and, if relevant, registered) have been explained.

## Contributor and guarantor information

CH, FS and KHW contributed to the planning, conduct and reporting of the work described in the article. All other authors contributed to the reporting of the work. As guarantor, KHW is responsible for the overall content of the article and attests that all listed authors meet authorship criteria and that no others meeting the criteria have been omitted.

## Copyright/license information

The Corresponding Author has the right to grant on behalf of all authors and does grant on behalf of all authors, a worldwide licence to the Publishers and its licensees in perpetuity, in all forms, formats and media (whether known now or created in the future), to i) publish, reproduce, distribute, display and store the Contribution, ii) translate the Contribution into other languages, create adaptations, reprints, include within collections and create summaries, extracts and/or, abstracts of the Contribution, iii) create any other derivative work(s) based on the Contribution, iv) to exploit all subsidiary rights in the Contribution, v) the inclusion of electronic links from the Contribution to third party material where-ever it may be located; and, vi) licence any third party to do any or all of the above.“

### Box 1 Mendelian randomization (MR) terminology and common methodology

#### Mendelian randomization (MR) methodological settings

##### One-sample MR or MR with individual-level data

MR analyses in which the associations between the instrumental variable (IV) and both the exposure and outcome are estimated from the same sample and individual-level data are used to derive the MR estimate.

##### Two-sample MR or MR with summary-level data

MR analyses in which the associations between the IV and both the exposure and outcome are generated from different (non-overlapping) samples and summary-level data are used to derive the MR estimate.

#### Most common MR methods used for main analyses

##### Two-stage least squares (TSLS)

Method implemented in a one-sample setting, where the first of two analytical stages involves estimating the association between the IV and continuous exposure through a linear regression model. The second stage then estimates the association between these genetically predicted exposure levels and the outcome using an appropriate regression model (i.e., linear or logistic regression, depending on the outcome variable). After adjusting standard errors to account for uncertainty in the estimates from the first stage, the TSLS method provides an estimate of the causal effect of the exposure on the outcome. The TSLS method can be applied with a continuous or categorical IV (e.g., a polygenic risk score or singular genotype, respectively).

##### Wald ratio

MR estimator for a single IV, where the effect estimate describing the association between the IV and the outcome is divided by the effect estimate describing the association between the IV and the exposure. The Wald ratio is equivalent to the TSLS method where there is one IV and can be used in either one- or two-sample settings and is, indeed, the basis of many two-sample MR methods.

##### Inverse variance weighted (IVW)

Most commonly applied method that combines Wald ratio estimates together in fixed effects meta-analysis, where the weight of each ratio is the inverse of the variance of the association between the IV and the outcome.

#### Sensitivity analyses

##### Overidentification or heterogeneity tests

Where there are multiple IVs, substantial differences (i.e., heterogeneity) across effect estimates may indicate that at least one IV is invalid. Overidentification tests (e.g., the Sargan test, traditionally used within one-sample MR analyses) or heterogeneity tests (e.g., Cochran’s Q, traditionally used within two-sample settings) can be used to test for the existence of heterogeneity between IVs and, therefore, the possibility that MR effect estimates may be biased due to violations of MR assumptions. However, if all IVs are biased in the same way, these tests will not be able to accurately identify such bias.

##### Pleiotropy-robust methods

Methods that test and adjust MR effect estimates for the presence of horizontal pleiotropy. For example, in one-sample settings, the Some Invalid Some Valid Instrumental Variable Estimation (SISVIVE), MR using G-Estimation under No Interaction with Unmeasured Selection (MR-GENIUS) and constrained IVs methods can be used. As most pleiotropy-robust methods have been historically developed for two-sample MR, there are many more tests and adjustments for pleiotropy with summary-level data including, most commonly, the weighted median, weighted mode and MR-Egger methods. Similarly, outlier-detection methods such as Mendelian Randomization Pleiotropy RESidual Sum and Outlier (MR-PRESSO) and Radial MR seek to remove outliers contributing high levels of heterogeneity to the MR effect estimate, which may indicate IV invalidity.

*For full details on MR terminology, methodology, assumptions and limitations, please see the MR Dictionary (*https://mr-dictionary.mrcieu.ac.uk/*) and Sanderson et al. Nature Reviews Methods Primers 2022*^1^.

### Box 2 Common reasons for studies being judged as “low quality”, “moderate quality” or “high quality” using the quality assessment tool

| QA item no. | QA item | Reasons for being judged “low quality” | Reasons for being judged “moderate quality” | Reasons for being judged “high quality” |
| --- | --- | --- | --- | --- |
| 1 | IV selection (1) | Using an inappropriate p-value for selecting IVs (e.g., a lenient p-value threshold $>5 \times 10^{-08}$), violating the first MR assumption and likely leading to the inclusion of SNPs that violated other MR assumptions. | Unclear what the p-value threshold for selecting IVs was or multiple p-value thresholds were used. | Used the most appropriate p-value threshold (e.g., the genome-wide significant p-value threshold from GWASs of $5 \times 10^{-08}$) for selecting instruments, ensuring the use of the most statistically robust instruments. |
| 2 | IV selection (2) | Including/excluding SNPs as IVs from primary analyses based on inappropriate criteria including lenient clumping methodology (e.g., not using an r-squared threshold of 0.001), selecting IVs based on F-statistics and excluding SNPs based on their associations with other phenotypes that could be on the causal pathway. | Unclear instrument selection criteria (e.g., not enough information was provided about the clumping strategy) or only one or two inappropriate criteria were used to select IVs (e.g., clumping threshold, F-statistics and phenotypic associations). | No studies had a clear enough strategy for instrument selection to be judged as high quality. What would have been required is clear and robust clumping strategy and an explicit statement indicating that F-statistics and phenotypic associations were not used for instrument selection. |
| 3 | Weak instrument bias | No studies presented results where the IV strength for all analyses undertaken was weak. However, it is worth noting that several studies presented many MR results, some of which had F-statistics $< 10$, indicating weak instrument bias. | Instrument strength (e.g., the F-statistic) was either not provided or discussed. In some cases, authors stated that the IVs were strong but did not provide statistical evidence for this. | Strength of the instruments used for MR analyses were explicitly described and statistically shown to be strong (e.g., with an F-statistic $> 10$). However, this was due to many studies explicitly removing SNPs that had an F-statistic $< 10$ before analyses were conducted, which is not best practice (and hence why many studies were not judged as “high quality” for the IV selection (2) item on the quality assessment tool). |
| 4 | Genetic confounding | Authors removed SNPs associated with individual-level confounders of the exposure-outcome association (usually before MR analyses) or showed that SNPs were associated with possible confounders and did not explore this further. | Authors either attempted to test for relationships with external variables that could induce bias via confounding (e.g., population-level confounders) but did not explore this in the MR analysis or this was not mentioned. | Authors explored population-level confounding (e.g., colocalization testing genetic confounding if only single SNPs were available) or explicitly showed that there were no associations between SNPs and individual-level confounders (as markers of population-level confounding). |
| 5 | Other<br>Confounding /<br>Collider bias | Data used for MR estimation explicitly adjusted for covariates that could induce bias (e.g., through collider bias or confounding) and inconsistencies in data structure. | Not clear whether covariates were added into the MR model (i.e., either directly or through inclusion in the GWAS sources) that could induce bias. | Data used for MR effect estimation did not adjust for any covariates that could induce bias, and any covariates included were similar across exposure and outcome data sources. |
| 6 | Additional direct effects between IV and outcome (exclusion restriction assumption) | Methods to test the robustness of results to violations of the third MR assumption were not undertaken. | Methods were not described transparently or clearly justified. | Methods were undertaken, well described and clearly justified. |
| 7 | Bias due to selection of participants | Not using appropriately homogenous samples when conducting two-sample MR analyses. | Information about population structure (usually indicated through genetic ancestry) or how contributing GWASs controlled for population structure (e.g., principal components) was not provided. | Data sources used for exposure or outcome information for two-sample MR analyses were from homogenous populations and/or weights for one-sample MR analyses were sourced from a population comparable to the data in which MR effects were estimated. |
| 8 | Reverse causation | Reverse causation was assessed; however, analyses were not accurately reported (e.g., conclusions indicated that there was evidence for an effect, but sensitivity analyses were inconsistent, indicating that there was limited robust evidence for an effect in the reverse direction). In some cases, results were not transparently reported as only p-values were presented. | Reverse causation was either not assessed or analyses were undertaken to test for reverse causation, but methodology was unclear, or results were not presented clearly or discussed. | Addressed reverse causation, where reverse MR analyses were explicitly undertaken and clearly presented and discussed. |
| 9 | Harmonization / preparation | Harmonization likely undertaken incorrectly. | Unclear whether the data had been harmonized appropriately. | Harmonization clearly described and appropriate. |
| 10 | Overestimation via sample overlap or overfitting | Explicit overlap in data sources contributing to MR analyses (i.e., samples used for exposure and outcome sources for two-sample MR analyses were not independent or GWAS used to generate weights for GRS for one-sample MR analyses was not independent of the data used to generate MR effect estimates). | Either not clear whether there was overlap between studies used in two-sample MR analyses or the source of weights for one-sample MR analyses was unclear. | Explicitly no overlap in data sources contributing to MR analyses. |
| 11 | Main analyses | No results were presented | Results were presented in | Results were presented and |
|  |  | in the text or only p-values were presented in the text, so no interpretation of effect estimates provided. | both text and tables, but the unit of interpretation or method used was unclear, with results presented often only based on reaching a specific p-value threshold. Where there was limited evidence of an effect, no results are presented. | interpreted clearly in the text and tables, with reference to units of the exposure and outcome. |
| 12 | Sensitivity analyses | Analyses testing the robustness of main effects to violations to MR assumptions (predominantly the “non horizontal pleiotropy” assumption) were not conducted or, where analyses had been conducted, they were either not reported in the text or explicitly misinterpreted. | Sensitivity analyses conducted and mostly reported but not entirely transparently, or no main result was found so sensitivity analyses were not undertaken. | Utilised sensitivity analyses to test for the robustness of results to violations of MR assumptions and transparently reported with comparison to main analyses. |
| 13 | Replication | No replication with an independent outcome dataset or replication effort was sought but this was not considered a formal replication (i.e., no independent data source was used). | Unclear whether efforts to replicate were legitimate (e.g., it was unclear what the replication dataset was) or validation analyses undertaken (e.g., different study design), to which MR estimates were compared. | Replication using appropriate methodology and external outcome data (for a two-sample MR analysis) or independent dataset (for one-sample MR analyses). |
| 14 | Data | Information about all data sources used in MR analyses was not clear. | Information about data sources was mostly clear, with some details missing about one of either the exposure or outcome data sources used. | Studies clearly referenced the data from which both the exposure and outcome were sourced were clear, and the information matched the referenced sources. |
| 15 | Exposure information | Neither exposure unit nor measurement clear. | Either exposure unit or measurement unclear. | Both exposure unit and measurement clear. |
| 16 | SNP information | Information about the number of SNPs used in MR analyses were not provided. | Insufficient information about how the number of SNPs used in analyses were attained. | Information about SNPs used in analyses are provided and clear why this number was used. |
| 17 | Outcome information | Neither outcome unit nor measurement clear. | Either outcome unit or measurement unclear. | Both outcome unit and measurement clear. |
| 18 | Reporting | Effect estimates not transparently or accurately presented, where conclusions are misleading, or no results presented at all. | Effect estimates presented are clear in terms of interpretation (where perhaps not all results presented), or all results are presented but interpretation (with consideration of main and sensitivity analyses) could have been clearer. | Results and effect estimates presented transparently, where main and sensitivity analyses have been considered appropriately when making causal inferences. |
| 19 | Data availability and reproducibility | Data used for analyses were not clear, analytical code was not provided, and methods were not | Data used for analyses were not clear (but provided) or were clear but analytical code was not provided (or | No study had entirely reproducible analyses. What would have been expected is that code and data were |
|  |  | transparently reported in a manner that was reproducible. | vice versa), and methods were not transparently reported in a manner that was reproducible. | provided and methods were transparently reported for full reproducibility. |

*GRS = genetic risk score; GWAS = genome-wide association study; IV = instrumental variable; MR = Mendelian* *randomization; QA = quality assessment; SNP = single nucleotide polymorphism. Most common reasons for a* *study being judged as “low quality”, “moderate quality” or “high quality” across the 19-item quality assessment* *(QA) tool.*

### Box 3 Best practices in undertaking Mendelian randomization analyses of microbial phenotypes

#### Best practices for undertaking MR analyses

##### P-value selection

To ensure the first MR assumption is valid, the IVs used must be robustly associated with the exposure, where the most robust threshold is the genome-wide p-value threshold (i.e., 5×10^−08^, adjusting for the number of independent genetic variants in typical GWASs, with an additional adjustment of the number of independent microbial traits being analysed). Ideally, the association between an IV and exposure would also be replicated across independent GWASs.

##### Other IV selection criteria

To ensure that the IVs used are independent (which prevents some bias due to violations of the second MR assumption but also to prevent overcounting any IV-specific estimate), robust clumping must be performed, where the traditional thresholds are an r-squared of <0.001 within a 10,000kb genomic window (or the explicit use of MR analyses that adjust for correlated SNPs). F-statistics should not be used to select IVs, as these are indicators of weak instrument bias when interpreting results. Ideally, SNPs should also not be excluded before any analyses due to associations with other phenotypes or indeed the outcome, as this may remove valid SNPs where associations reflect vertical pleiotropy. Where effect allele frequencies are available (or can be inferred), palindromic SNPs should not be removed unless they have an intermediate allele frequency (i.e., a MAF > 0.42). Equally, proxy SNPs (if available) could be used if palindromic SNPs are be removed.

##### GWAS data

GWASs used in a two-sample setting need to also be independent (and, ideally, if unavoidable, overlap should be calculated, with discussion of how much bias is likely induced) and, similarly, weights for one-sample MR analyses should be obtained from an independent data source than that used for undertaking MR analyses to avoid overfitting. Many GWASs adjust for study-specific covariates (e.g., batch effects and principal components), which is important for adjusting for population structure that may eventually bias MR effects due to violations of the second MR assumption. However, it is important not to use GWAS data that have adjusted for phenotypes that may be on the causal pathway in an MR analysis, as this can induce collider bias and, where exposure and outcome GWASs (i.e., for a two-sample MR analysis) have adjusted for different covariates, this can lead to samples that are not reflective of the same underlying population.

##### Combining data sources

SNPs being used as IVs for an exposure should not be sourced and combined across different exposure GWASs unless undertaking formal discovery and replication phases or a meta-analysis, as the SNP-exposure relationships may be reflective of different phenotypes with varying units meaning MR effect estimates are uninterpretable.

##### Harmonization

Details of the harmonization process need to be explicit, where, ideally, the number of SNPs lost at various stages of this process are reported for full transparency and reproducibility or, certainly, any assumptions being made (e.g., the GWASs were coded on the forward strand) and parameters being used (e.g., for proxy SNPs) are reported.

##### Testing the validity of the independence assumption

Whilst the second MR assumption cannot be tested statistically in practice, there are choices that can be made to reduce the likelihood that it is violated (e.g., using independent SNPs, use GWASs reflective of the same underlying population that adjust for population structure that could induce spurious associations between genetic variation and phenotypes). Testing for relationships between SNPs and individual-level confounding of the exposure and outcome (e.g., socioeconomic position) can be useful in identifying population-level confounding (i.e., which can lead to violations of the second MR assumption). However, this should be undertaken in sensitivity analyses exploring the causal effect estimates obtained (e.g., using multivariable MR or, if only single SNPs are available, methods like colocalization are useful), rather than simply removing SNPs that have associations with individual-level confounders.

##### Testing the validity of the exclusion restriction assumption

As sensitivity analyses to test robustness of main results, appropriate pleiotropy-robust methods and/or outlier detection tests with justification are useful to comprehensively understand the likely biases and causal nature of results. Where only single SNPs are available, testing the associations between SNPs and phenotypes (e.g., in a PheWAS) is useful; however, this does not allow the distinction between vertical or horizontal pleiotropy.

##### Reverse causation

Especially important in the context of microbiome MR studies, where SNPs may influence the microbial trait via the outcome of interest, reverse causality should be tested (e.g., with formal bi-directional MR analyses or, if only single SNPs are available, methods like colocalization are helpful).

##### Replication

Studies should strive for replication, whereby the same MR analyses are conducted in independent data sources (i.e., separate individual-level data for one-sample MR analyses or different outcome GWASs for two-sample MR analyses).

##### Reporting

To improve the reporting quality of MR results, there needs to be less reliance on p-values for interpretation and, instead, more focus on effect estimates and, especially when comparing sensitivity analyses, the consistency of effect estimates and their precision across methods in the context of power. Overall, there needs to be accurate and cautious interpretation of any estimates, especially in the context of the lack of biological understanding of the mechanisms by which human genetic variation influences the gut microbiome and the limited number of robust SNPs available for MR analyses.

##### Reproducibility

To ensure reproducibility of methods, accurate information on methods for main and sensitivity analyses (and their justification), data sources and participants, the number of SNPs utilised, harmonization

