## Supplementary Materials for "A systematic review of the applications of Mendelian randomization assessing the causal relevance of the gut microbiome in human health and disease"

- **Supplementary File 1** – Extracted data across 66 studies included
- **Supplementary File 2** – Quality assessment tool
- **Supplementary File 3** – Quality assessment results

*Search strategy*

OVID MEDLINE and EMBASE

1. Mendelian randomi#ation.hw,tw,kw,kf
2. genetic instrument?.hw,tw,kw,kf
3. (genetic variant* adj3 instrumental variable*).tw,kw,kf
4. gastrointestinal microbiome.sh
5. enteric microbial.hw,tw,kw,kf
6. (gut and (microbiome or microbiota or microflora or bacteria*)).tw,kw,kf
7. (intestinal and (microbiome or microbiota or microflora or bacteria*)).tw,kw,kf
8. (f#ecal and (microbiome or microbiota or microflora or bacteria*)).tw,kw,kf
9. or/1-3
10. or/4-8
11. 9 and 10
12. remove duplicates from 11

Fields: **hw** ([subject] heading word); **kf** (keyword heading word (MEDLINE)); **kw** (keyword (Embase); keyword heading (MEDLINE)); **tw** (text word).

Web of Science

(TI=(("Mendelian randomization" or "Mendelian randomisation" or "genetic instrument*" or ("genetic variant*" and "instrumental variable*")) AND (“gastrointestinal microbiome” or “enteric microbial” or (“gut” and (“microbiome” or “microbiota” or “microflora” or “bacteria*”)) or (“intestinal” and (“microbiome” or “microbiota” or “microflora” or “bacteria*”)) or (“fecal” and (“microbiome” or “microbiota” or “microflora” or “bacteria*”)) or (“faecal” and (“microbiome” or “microbiota” or “microflora” or “bacteria*”)))))

OR

(AB=(("Mendelian randomization" or "Mendelian randomisation" or "genetic instrument*" or ("genetic variant*" and "instrumental variable*")) AND (“gastrointestinal microbiome” or “enteric microbial” or (“gut” and (“microbiome” or “microbiota” or “microflora” or “bacteria*”)) or (“intestinal” and (“microbiome” or “microbiota” or “microflora” or “bacteria*”)) or (“fecal” and (“microbiome” or “microbiota” or “microflora” or “bacteria*”)) or (“faecal” and (“microbiome” or “microbiota” or “microflora” or “bacteria*”)))))

OR

(AK=(("Mendelian randomization" or "Mendelian randomisation" or "genetic instrument*" or ("genetic variant*" and "instrumental variable*")) AND (“gastrointestinal microbiome” or “enteric microbial” or (“gut” and (“microbiome” or “microbiota” or “microflora” or “bacteria*”)) or (“intestinal” and (“microbiome” or “microbiota” or “microflora” or “bacteria*”)) or (“fecal” and (“microbiome” or “microbiota” or “microflora” or “bacteria*”)) or (“faecal” and (“microbiome” or “microbiota” or “microflora” or “bacteria*”)))))

Fields: **TI**: title; **AB**: abstract; **AK**: author keyword;

bioRxiv and medRxiv

mendelian AND random AND (microbiome OR microflora OR bacteria OR bacterial OR microbial OR microbiota)

**Supplementary Table 1 – Characteristics of included studies**

| Review ID | First author | Year | Study designs | Exposure data sources | Broad outcome groups | Outcome data sources | Ref |
| --- | --- | --- | --- | --- | --- | --- | --- |
| 0635532754 | Mingyi Yang | 2022 | Two-sample | Hughes 2020 | Cardiovascular | UK Biobank | ^63^ |
| 1055369512 | Liling Lin | 2022 | Two-sample | Kurilshikov 2021 | Prescription drugs, Pain | Wu 2019, Johnston 2019 | ^75^ |
| 1108156353 | Linjing Zhang | 2022 | Two-sample | Wang 2016 | Brain | Nicolas 2018 | ^86^ |
| 122541860 | David A. Hughes | 2020 | Two-sample | Hughes 2020 (Current study) | Brain, Metabolic health, Autoimmunity | Jansen 2019, Locke 2015, Liu 2015, Ripke 2013, Morris 2012, Shungin 2015, Okada 2014, Simón-Sánchez 2009 | ^9^ |
| 1234567891 | C.A.F. Rivier | 2022 | Two-sample | Not reported | Cardiovascular | Unknown GWAS | ^55^ |
| 1286056190 | Min Chen | 2022 | Two-sample | Kurilshikov 2021 | Brain | Wray 2018 | ^94^ |
| 1308760143 | Qian Yang | 2018 | Two-sample | Turpin 2016, Mixture of mGWASs, Goodrich 2016 | Metabolic health, Cardiovascular | Willer 2013, Morris 2012, Deloukas 2013, Yang 2018 (Current study), Locke 2015, Dupuis 2010, Unknown GWAS, Nikpay 2015 | ^92^ |
| 1324207092 | Kun Xiang | 2022 | Two-sample | Kurilshikov 2021 | Metabolic health | Spracklen 2020, Xue 2018 | ^80^ |
| 1519899723 | Maxim B. Freidin | 2021 | One-sample | Freidin 2021 (Current study) | Pain | Freidin 2021 (Current study) | ^46^ |
| 1599546107 | Xiaolin Yang | 2022 | Two-sample | Kurilshikov 2021 | Nutrition | Revez 2020 | ^70^ |
| 1795779689 | Yuzhen Ouyang | 2022 | Two-sample | Kurilshikov 2021 | Brain | ILAE 2018 | ^53^ |
| 1828507192 | Esteban A. Lopera-Maya | 2022 | Two-sample | Lopera-Maya 2022 (Current study) | Behaviour, Brain, Immune system, Nutrition, Metabolic health, Autoimmunity, Cardiovascular, Bone | UK Biobank, Lambert 2013, Boraska 2014, Moffatt 2010, Locke 2015, Dubois 2010, Schunkert 2011, Liu 2015, Paternoster 2015, Kottgen 2013, Malik 2018, Willer 2013, Sawcer 2011, Simón-Sánchez 2009, Okada 2014, Hom 2009, Morris 2012, Shungin 2015 | ^95^ |
| 1915685263 | Young Ho Lee | 2020 | Two-sample | Not reported | Autoimmunity | Unknown GWAS | ^51^ |
| 1977124126 | Jing Ning | 2022 | Two-sample | Kurilshikov 2021 | Brain | Lambert 2013, Nalls 2019, Nicolas 2018 | ^38^ |
| 2044689642 | Serena Sanna | 2019 | Two-sample | Sanna 2019 (Current study) | Metabolic health | Prokopenko 2014, Shungin 2015, Saxena 2010, Dupuis 2010, Manning 2012, Soranzo 2010, Locke 2015, Scott 2017, Strawbridge 2011, UK Biobank | ^24^ |
| 2093521025 | Qian Xu | 2021 | Two-sample | Goodrich 2016 | Metabolic health | Xu 2021 (Current study) | ^91^ |
| 2136589463 | Lianmin Chen | 2022 | Two-sample | Chen 2022 (Current study) | Metabolic health | Chen 2022 (Current study) | ^72^ |
| 2179267930 | Xiaomin Liu | 2022 | One-sample, Two-sample | Liu 2022 (Current study) | Metabolic health, Cardiovascular, Cancer, Lung, Skin, Immune system, Prescription drugs, Kidney, Nutrition, Liver, Sexual and reproductive health, Eyes, Mouth, Brain, Bone | Liu 2022 (Current study), Ishigaki 2020 | ^26^ |
| 2238214110 | Xin Qi | 2021 | Two-sample | Mixture of mGWASs | Brain, Liver, Prescription drugs, Nervous system, Behaviour, Medical procedure, Gastrointestinal tract, Inflammation, Pain, Cardiovascular, Bladder, Eyes, Pancreas, Cancer, Sexual and reproductive health | Unknown GWAS, UK Biobank | ^54^ |
| 2300517094 | Wen-Di Shen | 2022 | Two-sample | Shen 2022 (Current study) | Metabolic health | Shen 2022 (Current study) | ^78^ |
| 23119582 | Nicholas Harvey | 2021 | Two-sample | Kurilshikov 2021 | Autoimmunity | Unknown GWAS | ^49^ |
| 2380923650 | Lei Hou | 2022 | Two-sample | Hughes 2020 | Behaviour, Nutrition, Longevity | Cole 2020, Codd 2014 | ^74^ |
| 2383481240 | Kangcheng Liu | 2022 | Two-sample | Kurilshikov 2021, Goodrich 2016 | Eyes | Kurki 2022 | ^87^ |
| 2419487002 | X.-H. Yu | 2021 | Two-sample | Kurilshikov 2021 | Bone | Tachmazidou 2019 | ^57^ |
| 2547037187 | Yuxuan Zhang | 2022 | Two-sample | Kurilshikov 2021 | Cardiovascular | Roselli 2018, Harst 2018, Nikpay 2015, Shah 2020, Malik 2018 | ^89^ |
| 261727485 | Tim Robinson | 2021 | Two-sample | Not reported | Cancer | Unknown GWAS | ^61^ |
| 2634842805 | Chuang Li | 2022 | Two-sample | Kurilshikov 2021 | Sexual and reproductive health | FinnGen Release 5 (2020) | ^65^ |
| 2662850683 | Alexander Kurilshikov | 2021 | Two-sample | Kurilshikov 2021 (Current study) | Brain, Immune system, Metabolic health, Cardiovascular, Autoimmunity, Bone, Liver, Behaviour, Nutrition | Unknown GWAS | ^10^ |
| 2738712179 | Dan He | 2022 | Two-sample | Hughes 2020 | Longevity | Zenin 2019, Timmers 2019, Deelan 2019, Pilling 2016 | ^96^ |
| 289377291 | S. L. Clarke | 2021 | Two-sample | Not reported | Autoimmunity | Unknown GWAS | ^59^ |
| 2918940101 | Charlie Hatcher | 2022 | Two-sample | Hughes 2020 | Cancer | Hatcher 2022 (Current study) | ^98^ |
| 2934159401 | Kangcheng Liu | 2022 | Two-sample | Kurilshikov 2021, Goodrich 2016 | Inflammation | FinnGen Release 7 (2022), Jiang 2021 | ^25^ |
| 3032725182 | Jing-Jing Ni | 2021 | Two-sample | Goodrich 2016, Bonder 2016 | Bone | Morris 2018 | ^28^ |
| 3113131015 | Huaqiang Zhou | 2020 | Two-sample | Mixture of mGWASs | Cancer | Wang 2014 | ^93^ |
| 3132308026 | Hui-Min Liu | 2021 | Unclear | Liu 2021 (Current study) | Metabolic health, Cardiovascular | Liu 2021 (Current study) | ^22^ |
| 3148374251 | Eva M. Asensio | 2022 | Two-sample | Qin 2022 | Metabolic health | Asensio 2022 (Current study) | ^82^ |
| 3184652665 | Yuija Lu | 2023 | Two-sample | Sanna 2019 | Cancer | Huyghe 2019 | ^62^ |
| 3191171275 | Sergio Andreu-Sánchez | 2022 | Two-sample | Kurilshikov 2021 | Metabolic health | Andreu-Sánchez 2022 | ^47^ |
| 3272561227 | Zi-Jia Zhang | 2021 | Two-sample | Goodrich 2016 | Autoimmunity | Unknown GWAS | ^29^ |
| 3278202937 | Malte Christoph Rühlemann | 2021 | Two-sample | Rühlemann 2021 (Current study) | Brain, Immune system, Autoimmunity, Metabolic health, Kidney, Cardiovascular, Cancer, Bone, Gastrointestinal tract | Unknown GWAS | ^23^ |
| 3286259646 | Jing-Jing Ni | 2022 | Two-sample | Kurilshikov 2021 | Brain | Grove 2019, Stahl 2019, PGC 2014, Yu 2019, van den Berg 2016, Nagel 2018, Erlangsen 2020, IOCDF-GC+OCGAS 2018, Demontis 2019 | ^68^ |
| 3549710879 | Zhenhuang Zhuang | 2022 | Two-sample | Wang 2016 | Autoimmunity | Liu 2015 | ^90^ |
| 3678116995 | Pengsheng Li | 2022 | Two-sample | Kurilshikov 2021 | Sexual and reproductive health | Kurki 2022 | ^76^ |
| 3760378084 | Bin Liu | 2022 | Two-sample | Kurilshikov 2021 | Autoimmunity | Liu 2015 | ^71^ |
| 3836325935 | Jun Inamo | 2021 | Two-sample | Mixture of mGWASs | Autoimmunity | Okada 2014 | ^50^ |
| 4090935045 | Shu-Hui Xie | 2021 | Two-sample | Not reported | Metabolic health | Francis 2021 | ^43^ |
| 4163561095 | Djawad Radjabzadeh | 2022 | Two-sample | Kurilshikov 2021 | Brain | Howard 2019 | ^60^ |
| 4199071353 | Zhao Yang | 2022 | Two-sample | Kurilshikov 2021 | Oesophagus | Ong 2021 | ^77^ |
| 4251029218 | Jing-Jing Ni | 2022 | Two-sample | Goodrich 2016, Wang 2016 | Cancer | Zhou 2018 | ^67^ |
| 4286528530 | Qiang Luo | 2022 | Two-sample | Kurilshikov 2021 | Cardiovascular, Kidney, Metabolic health | Nielson 2018, Pattaro 2016, Harst 2018, Xue 2018, Evangelou 2018, Sakaue 2021, Shah 2020, Nikpay 2015 | ^52^ |
| 4290593439 | Hilde E. Groot | 2020 | Two-sample | Scepanovic 2019, Wang 2016, Bonder 2016, Goodrich 2016, Turpin 2016, Unclear mixture of mGWASs | Cardiovascular, Immune system, Metabolic health, Lung, Behaviour | Groot 2020 (Current study) | ^83^ |
| 505573255 | Zhenhuang Zhuang | 2020 | Two-sample | Wang 2016 | Brain | Lambert 2013, Wray 2018, Ripke 2013 | ^88^ |
| 557846834 | Yilin Chen | 2020 | Two-sample | Not reported | Cardiovascular | Unknown GWAS | ^58^ |
| 560890194 | Ulrika Boulund | 2022 | Two-sample | Boulund 2022 (Current study) | Metabolic health, Cardiovascular | Pan UK Biobank GWAS | ^48^ |
| 568199176 | Wan-Qiang Lv | 2021 | One-sample, Two-sample | Lv 2021 (Current study), Sanna 2019 | Metabolic health | Lv 2021 (Current study), Zillikens 2017 | ^30^ |
| 601575515 | Iraia García-Santisteban | 2020 | Two-sample | Bonder 2016 | Autoimmunity | Dubois 2010 | ^31^ |
| 707846668 | Qian Xu | 2021 | Two-sample | Kurilshikov 2021 | Autoimmunity | Bentham 2015, Okada 2014, Patsopoulos 2020, de Lange 2017, Onengut-Gumuscu 2015, Trynka 2011, Julia 2018, UK Biobank, Dubois 2010 | ^27^ |
| 718621312 | Youwen Qin | 2022 | Two-sample | Qin 2022 (Current study) | Brain, Cancer, Kidney, Immune system, Bone | Lambert 2013, van Rheenen 2016, Duncan 2017, Unknown GWAS, Michailidou 2017, Pattaro 2016, Paternoster 2015, Kottgen 2013, Zeggini 2012, Wang 2015, Wray 2018, Albagha 2011, Ripke 2014 | ^69^ |
| 776143774 | Kun Xiang | 2021 | Two-sample | Kurilshikov 2021 | Autoimmunity | Bentham 2015 | ^85^ |
| 878349061 | C. Wijmenga | 2019 | Unknown | Not reported | Metabolic health | Unknown GWAS | ^44^ |
| 880716487 | Wenchuan Zhang | 2022 | Two-sample | Kurilshikov 2021 | Metabolic health | Folkersen 2020 | ^45^ |
| 914635920 | Fengzhe Xu | 2020 | Two-sample | Xu 2020 (Current study) | Metabolic health, Behaviour, Cardiovascular, Kidney, Cancer, Autoimmunity | Japanese Biobank | ^73^ |
| 95001313 | Jun Ma | 2022 | Two-sample | Rühlemann 2021 | Cancer | Jiang 2021 | ^64^ |
| 950921645 | Young Ho Lee | 2021 | Two-sample | Mixture of mGWASs | Bone | Zeggini 2012 | ^66^ |
| 978492202 | P. Tejera | 2021 | Two-sample | Not reported | Respiratory disorders | Unknown GWAS | ^56^ |
| Other sources | Eloi Gagnon | 2023 | Two-sample | Mixture of mGWASs | Metabolic health, Cardiovascular, Kidney, Brain, Longevity, Liver, Bone | UK Biobank, Evangelou 2018, Stanzick 2021, Lagou 2021, Richardson 2020, Jansen 2019, Wuttke 2019, Harst 2018, Howard 2019, Deelan 2019, Malik 2018, Ghodsian 2021, Timmers 2019, Mahajan 2018 | ^79^ |

*GWAS = genome-wide association study (GWAS); ID = identification number; MR = Mendelian randomization; Ref = reference. Brief information on all 66 studies (with full details presented in Supplementary File 1), including the identification number used in the review, first author, year of publication or availability of pre-print or conference abstract, study design (e.g., where either one- or two-sample Mendelian randomization (MR) was used, if described), what data sources were used for the exposure, what types of outcome were analysed and where the outcome data were sourced.*

**Supplementary Figure 1** **– Flow chart for deciding if results can be meta-analysed**

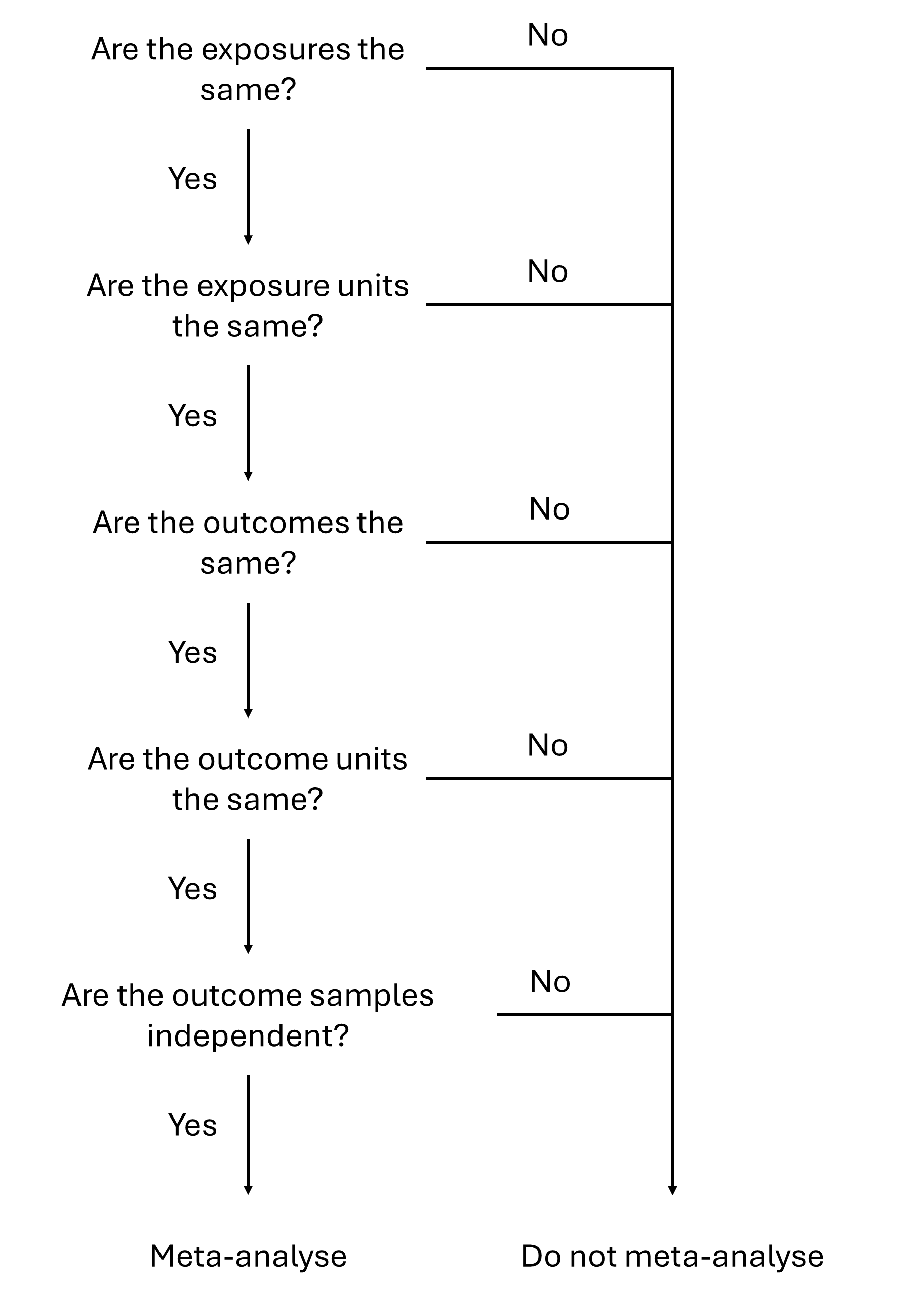

*To establish whether any estimates across studies were comparable and could be included in a meta-analysis, the estimates must represent the effect of the same exposure (i.e., name, definition and unit) on the same outcome (i.e., name, definition and unit). Whilst the source from which the exposure data were obtained could be the same across estimates, the source from which the outcome data were obtained must be independent across estimates. If all described criteria were met, the estimates could be meta-analysed; otherwise, they could not be meta-analysed.*

**Supplementary Figure 2 – Comparison of studies using the same data and analysis method**

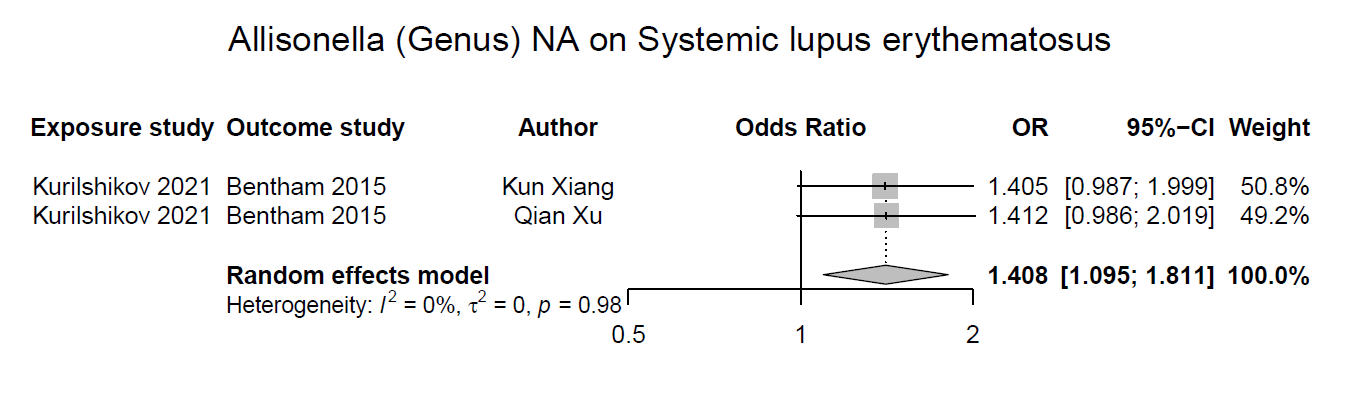

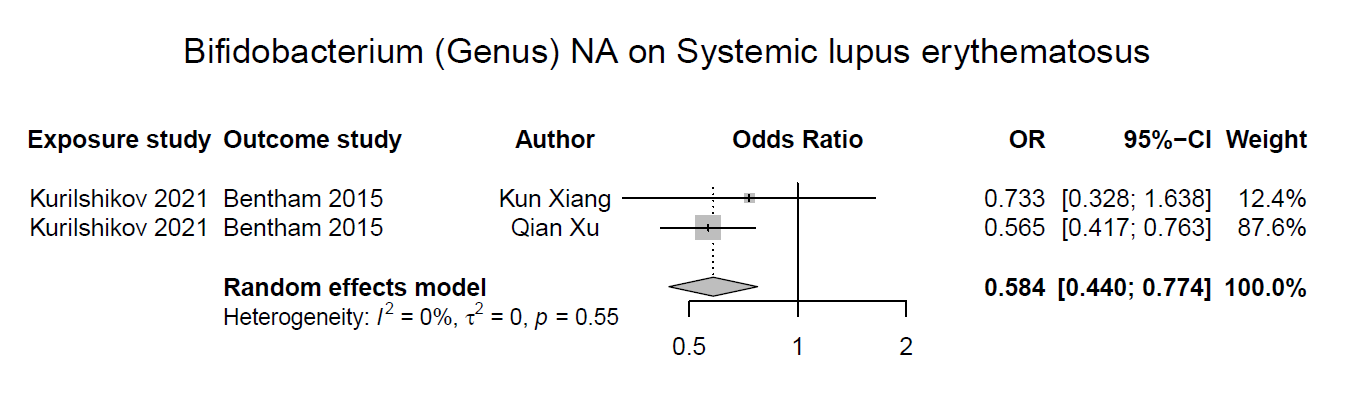
*A direct comparison was made where studies had explicitly undertaken the same analysis (i.e., the same exposure and unit, same outcome and unit, same data and same analysis method).*
